# A liquid biopsy-centered, pan-cancer, open next generation sequencing panel to support clinical decision-making (LION panel)

**DOI:** 10.64898/2026.06.05.26354976

**Authors:** Sabrina Feierabend, Axel Künstner, Michael Forster, Theresa Helbing, Niklas Gebauer, Timo Gemoll, Franziska Axt, Subbaiah Chary Nimmagadda, Lavanya Ranganathan, Johanna Schwandt, Marilena Heber, Silke Szymczak, Ina Hohensee, Stephanie M. J. Fliedner, Florian Scherer, Martina Oberländer, Stefanie Derer-Petersen, Hauke Busch, Nikolas von Bubnoff, Eva Dazert

**Author notes:** **Corresponding authors:** Axel Künstner, PhD, Medical Systems Biology Group, Institute of Experimental Dermatology, University of Luebeck, Ratzeburger Allee 160, 23562 Luebeck, Germany, Dr. Eva Dazert-Klebsattel, Klinik für Hämatologie und Onkologie, Biomedizinische Forschung, Ratzeburger Allee 160, Haus 67, 23562 Luebeck. Equal contribution and shared first authorship (Sabrina Feierabend, Axel Künstner).

## Abstract

Cancer treatment has shifted toward personalized therapy based on molecular profiling, particularly in advanced disease. Existing circulating tumor DNA panels are often broad, generating many non-actionable variants and incurring costs that limit routine use in molecular tumor boards. We developed and validated a manufacturer-independent, 109-gene liquid biopsy-centered pan-cancer open next generation sequencing panel (LION panel), combined with an in-house bioinformatic pipeline to support clinical decision-making.

A total of 87 samples were analyzed, including 17 reference samples, 21 healthy blood donor controls, and 49 patient samples including nine tumor entities.

The LION panel achieved 92% sensitivity and 99% specificity in reference samples, with high concordance to digital droplet PCR (r = 0.99). It detected variant allele frequencies as low as 0.05% (tumor-informed) and 0.5% (tumor-uninformed). Clinical concordance reached 82% with blood-based digital droplet PCR and 75% with whole exome tissue sequencing. In representative cases, variant dynamics correlated with disease progression and revealed additional targetable variants.

Overall, the LION panel supports clinical decision-making by enabling identification of targetable variants, disease monitoring, and detection of treatment resistance, particularly when tumor tissue is unavailable.

## 1 Introduction

The treatment of cancer patients has evolved from a one-size-fits-all approach towards personalized therapy. This precision oncology approach comprises specific targeted therapies based on molecular diagnostics of the tumor DNA or RNA (1). A large prospective observational study by Horak et al. has demonstrated that extensive molecular analysis supports clinical decision-making, particularly in patients with rare or advanced cancer and can lead to improved outcomes (2). However, treatment resistance remains a major challenge in targeted cancer therapy (1).

To identify evidence-based treatment options for various tumor diseases in patients with advanced cancer, molecular tumor boards (MTBs) have been established. These multidisciplinary research teams are essential as molecular data become increasingly complex, requiring expertise from multiple disciplines, while the number of targeted treatment options continues to expand (3,4). Molecular analyses are not limited to tumor tissue biopsies. They can also be performed on liquid biopsies, although their clinical implementation is still evolving (5).

Tumors typically receive rich vascularization, allowing them to release not only cells into the bloodstream but also fragments of cell-free DNA, known as circulating tumor DNA (ctDNA). This release occurs through processes such as apoptosis, necrosis or secretion by tumor cells, and liquid biopsy enables the non-invasive analysis of ctDNA from peripheral blood samples and the detection of tumor-specific genetic alterations (6). Compared with tissue biopsy collection, a liquid biopsy is associated with reduced procedural risk, enables repeated sampling and reflects the actual tumor heterogeneity as opposed to the locally restricted information of single tissue biopsies. Therefore, liquid biopsy represents a promising tool for longitudinal disease monitoring, including detection of progression, recurrence, treatment resistance, and minimal residual disease (MRD) (7).

For MRD monitoring and early detection of progression and treatment resistance, highly sensitive methods capable of identifying variants with a low variant allele frequency (VAF) in ctDNA are required (5). Current regulatory requirements, including the German medical laboratory quality assurance guideline (Rili-BAEK), mandate validation of next generation sequencing (NGS)-based molecular diagnostics with a lower limit of detection of at least 0.5% VAF for routine clinical application (8). However, variants below this threshold may carry biological or clinical relevance, underscoring the need for approaches with sensitivity beyond standard regulatory requirements. Targeted NGS panels offer a practical balance between analytical sensitivity, cost, and clinical turnaround time, making them well-suited for routine liquid biopsy workflows in MTBs.

Several technologies are available for ctDNA analysis, including digital droplet PCR (ddPCR) and NGS-based approaches, such as whole exome sequencing (WES) and targeted NGS. WES enables the analysis of all protein-coding regions of the genome, while its sensitivity for ctDNA detection is often limited by comparatively low sequencing depth and associated costs (9). In contrast, ddPCR offers very high analytical sensitivity, but is restricted to the detection of predefined mutations (10,11). Targeted NGS approaches enable parallel analysis of multiple alterations without prior knowledge of specific variants and can achieve high sensitivity depending on panel design and sequencing depth (12,13). Targeted NGS panels focusing on clinically relevant genomic regions have been shown to improve clinical decision-making and early detection of disease progression in several tumor entities, including non-small-cell lung cancer (NSCLC) (14).

Clinically used NGS panels are often designed for detecting single nucleotide variants (SNVs) and small insertions or deletions (indels) as well as gene fusions and copy number alterations (CNAs) in specific cancer-related genes. The spectrum of cancer-related genomic analysis and genotyping can be extended to include the analysis of genes associated with clonal hematopoiesis of indeterminate potential (CHIP) (15,16).

CHIP is characterized by somatic mutations in genes of peripheral blood cells associated with myeloid blood cancers in the absence of a hematological disorder. CHIP prevalence increases with age and is associated with elevated risks of hematologic cancers, coronary heart disease and overall mortality (17). Distinguishing CHIP-derived variants from tumor-specific mutations is therefore essential in ctDNA analysis. Although CHIP has emerged as potential prognostic factor, its relevance in solid tumors remains under investigation (18).

Gene fusions contribute to cancer development by introducing loss or gain of function effects or altered gene expression. Since fusion breakpoints frequently occur within intronic regions, detection is often more reliable at the RNA level than at the DNA level. Despite increasing clinical relevance, implementation of routine fusion diagnostics is still evolving. Notably, targetable fusions, including ROS1 fusions in NSCLC responsive to crizotinib and tumor-agnostic NTRK fusions, underscore the clinical importance of fusion detection in precision oncology (19–22).

Commercially available comprehensive sequencing panels are often not optimized for routine MTB workflows, as they may generate large numbers of clinically non-actionable mutations while increasing analytical complexity and costs (23). Therefore, we developed and validated a customized, manufacturer-independent 109-gene NGS panel together with an in-house bioinformatic pipeline to support clinical decision-making specifically based on liquid biopsy-derived ctDNA within the MTB (LION panel and pipeline).

We chose the 109 top cancer genes contained in the LION panel due to 1) their importance as main cancer driver genes covering the most important entities such as colorectal cancer and lung cancer (69% of the LION panels size), 2) their involvement in cancer drug resistance (18%), 3) their role in important cancer-related pathways such as CHIP (7%), 4) their aberration as fusion genes (4%) and finally their involvement in NGS quality control-related parameters such as harboring patient genotyping single nucleotide polymorphisms (SNPs).

We aimed to establish a sensitive and clinically applicable, freely available workflow for the detection of targetable mutations and recurrence and resistance development in a wide range of cancer entities, particularly in cases where the tumor is inoperable or tumor tissue is unavailable or of poor quality.

## 2 Materials and Methods

### 2.1 Custom NGS panel design

A customized hybrid-capture NGS panel (LION panel) was designed using SureDesign (Agilent Technologies, Santa Clara, CA, USA). The human reference genome GRCh38 was used for probe design. The panel targets 109 genes, covering 500 kb of coding sequence to ensure high coverage depth while maintaining feasibility for routine diagnostics. Gene selection aimed to cover multiple tumor entities and included major cancer drivers, genes associated with therapy resistance and recurrent oncogenic fusions. Genes frequently mutated in CHIP were included to facilitate discrimination between tumor variants and age-related hematopoietic mutations. All coding exons of the selected genes were included in the design. The clinically relevant hotspot regions were enriched to achieve approximately 2-fold greater sequencing depth compared to the non-hotspot regions. For the detection of gene fusions, a DNA-based capture strategy was applied targeting recurrent breakpoint-associated intronic regions and commonly involved exons as described by Seki et al. including the genes *ALK*, *BCR*, *BRAF*, *FGFR2*, *FGFR3*, *NTRK1*, *RET* and *ROS1* (24). Previously described sample-tracking single nucleotide polymorphisms (SNPs) according to Pengelly and colleagues were incorporated into the panel to enable sample identification and verification and detection of potential sample mix-ups (25,26). An overview of the panel design is shown in Fig. 1A and a complete gene list including annotated hotspot regions is provided in Supplementary Table S1.

**Figure 1.**
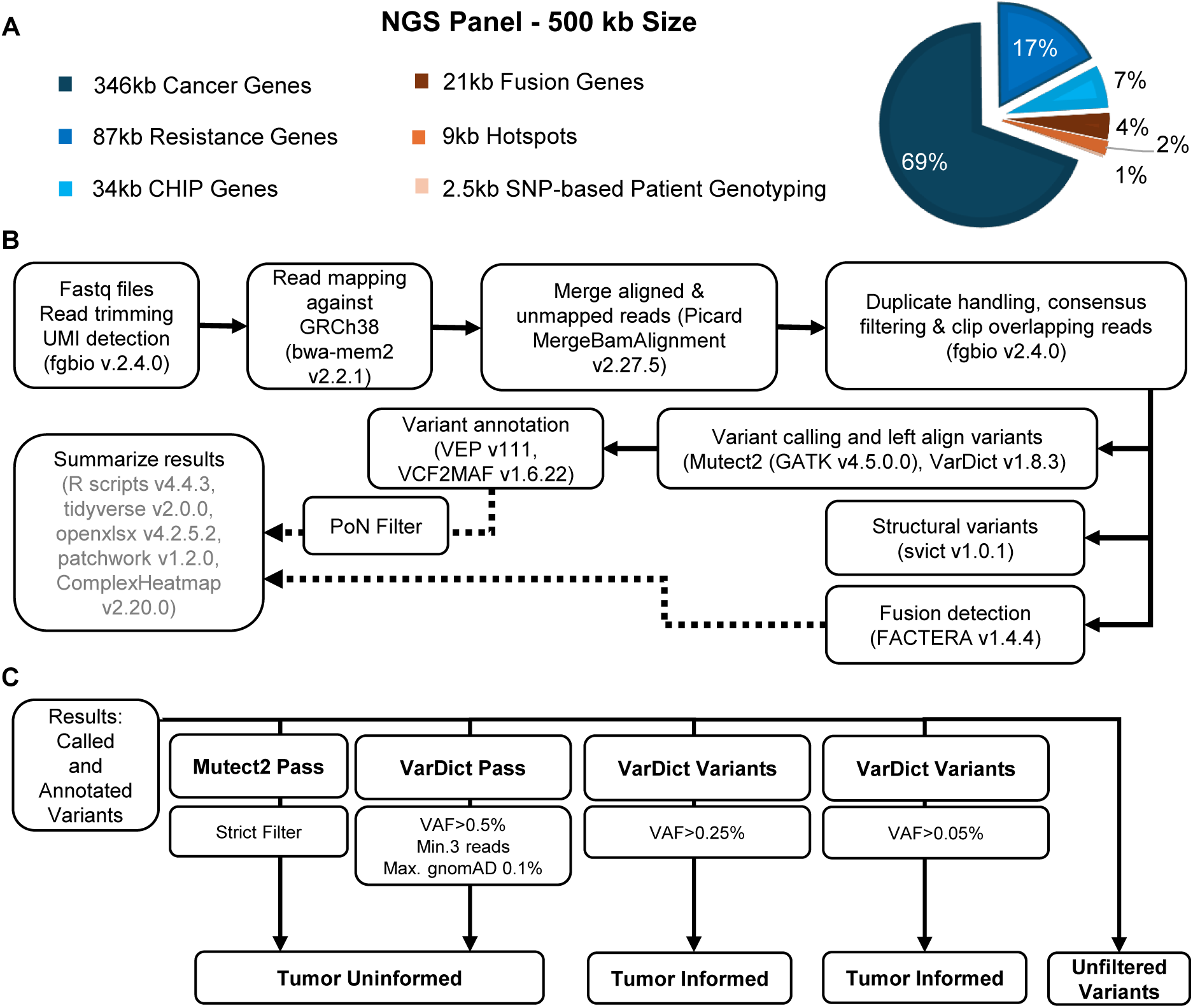
Overview of the LION NGS panel characteristics, bioinformatic and analysis workflow. (A) The pie chart illustrates the size distribution within the NGS panel based on clinically relevant gene categories. (B) Overview of the bioinformatic workflow used to generate summarized results from Illumina sequencing data. See Supplementary Figure S1 for additional details of the panel of normal (PoN) filter. (C) Overview of further data analysis of the summarized sequencing results. There are two different variant-callers: Mutect2 and VarDict, as well as specially adapted filter steps. Depending on the level at which the variants pass the filters, they are considered true positive if they were not previously known (tumor-uninformed) or if they were previously known (tumor-informed). CHIP, clonal hematopoiesis of indeterminate potential; gnomAD, genome aggregation database; KB, kilobase; Max., maximum; Min., minimum; NGS, next-generation sequencing; PON, panel of normal; SNP, single nucleotide polymorphism; VAF, variant allele frequency.

In detail, the 500kb of panel size comprises 346kb containing the most important cancer driver genes, such as kinases which are targetable, 87kb containing therapy resistance genes, 34kb containing genes involved in clonal hematopoiesis (CHIP), 21kb containing fusion genes, 9kb containing extra probes designed to cover the hotspots of the most important cancer driver genes (e.g. BRAF V600E, KRAS G12D) and finally 2.5kb containing probes covering single nucleotide polymorphisms (SNPs) used for patient genotyping as shown by Pengelly et al (25,26).

The final list is a selection of 109 genes that were included due to their presence in one or more of the following sources:

1. in the TruSight Oncology (TSO500) NGS panel (Illumina)
2. in the Ion AmpliSeq Comprehensive Cancer NGS panel (Thermo Fisher Scientific) and Cancer Hostpot NGS panel (Illumina)
3. in the Agilent SureSelect Cancer All-In-One (AIO) NGS panel (Agilent)
4. in the Nationales Netzwerk Genommedizin (nNGM) 2.0 NGS panel
5. in the Avenio ctDNA target NGS panel (Roche)
6. in the Oncomine™ Pan-Cancer Cell-Free Assay NGS panel (ThermoFisher)
7. in the IntOGenes platform Driver Genes selection for COREAD (Colon and Rectum adenocarcinoma)
8. in the non-small cell lung cancer (NSCLC)-specific NGS panel published by our group (14)
9. as cancer driver genes in Multiple myeloma (27–29) and Colorectal Cancer (30,31)
10. as therapy resistance genes
11. as CHIP genes (32)
12. as internal suggestions for cancer relevant genes of the University Hospital Schleswig-Holstein (UKSH), Campus Luebeck and Kiel
13. as fusion gene breakpoint regions as shown by Seki et al (24)
14. as quality control SNPs for patient genotyping (Pengelly SNPs) (25,26)

### 2.2. Reference samples and study cohort

The sample cohort analyzed in this study comprised positive controls (reference cfDNA samples), negative controls (plasma from healthy blood donors) and plasma/buffy coat samples from patients with various entities. Reference cfDNA samples (Seraseq ctDNA v2 Mutation Mix, HiSS Diagnostics GmbH, Freiburg, Germany), harboring 40 known mutations at low VAFs (0.25-1.0%), were used as positive controls to explore sensitivity at different cfDNA input amounts. Three reference materials were included, each harboring the same set of 40 reference mutations at VAFs of 0.25%, 0.5%, or 1%. These three different references were each measured each six times: twice with a DNA input of 50ng, twice with 25ng DNA input and twice with 10ng DNA input. The only exception was the 0.25% VAF reference at 10ng DNA input, for which only a single replicate was available. Consequently, a total of 17 technical replicates of commercial cfDNA references were analyzed. These reference samples were used for bioinformatic workflow optimization and analytical performance assessment. The 21 negative retrospective control plasma samples were measured from 21 different healthy blood donors who donated blood at the University Medical Center Schleswig-Holstein (UKSH) in Luebeck. These blood donor plasma samples were used to estimate and filter out false-positive mutations. Finally, to validate the panel for use in MTBs, an initial cohort comprising 85 samples from 71 patients admitted to the UKSH Luebeck or Kiel was retrospectively assembled. Following quality control and inclusion filtering, 49 clinical samples from 42 patients were retained for validation analyses. Overall, 17 reference samples, 21 healthy blood donor samples and 85 patient samples were screened in the initial study set (n=123). The final analysis set comprised 87 samples, including 17 reference samples, 21 healthy blood donor samples and 49 patient samples.

### 2.3 Ethics statement

Human samples included in this study originated from different cohorts and biobanking projects that were covered by separate ethics approvals. The respective study protocols were approved by the Ethics Committee of the University of Luebeck (2023-315; 2024-392_1; 19-377; 18-311; 16-282; 2023-577), the Ethics Committee of Kiel University (D 431/06), and the Institutional Review Board of the University Medical Center of Schleswig-Holstein (2022–634). The study was conducted in accordance with the Declaration of Helsinki.

### 2.4 Sample processing and DNA isolation

Peripheral blood samples were collected in EDTA (Sarstedt, Nümbrecht, Germany) or cell-free DNA-BCT® CE tubes (Streck, La Vista, USA) and processed according to the manufacturer’s instructions as detailed in the Supplementary Methods. Plasma and buffy coat were isolated by centrifugation and stored at -80 °C until further use. Cell-free DNA (cfDNA) was isolated from plasma using QIAmp® Circulating Nucleic Acid Kit (Qiagen, Hilden, Germany) or the EZ2 Connect with the EZ 1 & 2 ccfDNA Kit (Qiagen, Hilden, Germany), while gDNA from buffy coat and FFPE tissue was isolated using respective kits according to the manufacturer’s instructions (see Supplementary Methods). DNA concentration was quantified using a Qubit 4 Fluorometer (Thermo Fisher Scientific, Waltham, USA), and isolated DNA was stored at -20 °C until further use.

### 2.5 Library preparation and sequencing

Libraries were prepared using the SureSelect XT HS2 DNA Target Enrichment Kit (Agilent Technologies, Santa Clara, USA) according to the manufacturer’s instructions, initially manually and subsequently on the Magnis Dx NGS Prep System (Agilent Technologies, Santa Clara, USA), as detailed in the Supplementary Methods. Molecular-barcoded adapters (MBCs) containing unique molecular identifiers (UMIs) were used for duplicate-aware error correction. Final library quality was assessed using the Agilent 4150 TapeStation Instrument (Agilent Technologies, Santa Clara, USA) and the Qubit 4 Fluorometer (Thermo Fisher Scientific, Waltham, USA) (see Supplementary Methods). Libraries were multiplexed and sequenced on the NovaSeq6000 platform (Illumina, San Diego, USA) using paired-end 2 x 151 bp reads. Sequencing targeted a mean on-target coverage after bioinformatic error-correction of approximately 500x for FFPE and 3000x for liquid biopsy samples, as recommended by the library kit manufacturer.

### 2.6 Bioinformatic analysis and variant classification

Sequencing data were analyzed using a customized bioinformatic workflow (LION pipeline) (Fig.1B-C, (33)). Paired-end FASTQ reads were processed within a liquid biopsy-optimized pipeline. UMIs were extracted using fgbio (v2.4.0), followed by quality filtering with fastp (v0.24.0) (34). Reads were aligned to the human reference genome (GRCh38) using bwa-mem2 (v2.2.1) (35). Error correction included merging of aligned and unmapped reads using Picard (36) MergeBamAlignment (v2.27.5) to retain UMI tags, as well as PCR duplicate removal using Picard MarkDuplicates for internal quality control. Further error correction was achieved by grouping reads with identical UMIs using fgbio’s GroupReadsByUmi (paired strategy), followed by consensus calling with CallMolecularConsensusReads. These consensus reads were realigned to the reference genome and filtered with FilterConsensusReads to remove low-quality consensus families (minimum size of 2).

Overlapping read pairs were clipped using fgbio’s ClipBam to prevent double counting of variants. Somatic variant calling and left alignment of variants were conducted using VarDict (v1.8.3) (37) and Mutect2 (38). Variants were annotated using VEP (v111) (39) and converted to MAF format using VCF2MAF (v1.6.22). Fusion detection targeting chromosomal rearrangements within the panel design was performed using FACTERA (v1.4.4) (40).

Structural variant detection using svict (v1.0.1) (41) was implemented within the workflow but was not included in downstream analyses.

To reduce technical and population-specific background variants, a panel of normal (PoN) filter was applied based on sequencing data from 21 healthy blood donors. For inclusion in the PoN, a variant had to be detected in ≥ two healthy blood donor samples to avoid single-sample artefacts with a minimum of three reads in each sample after UMI-based error-correction. A variant was classified as recurrent background artefact if it was detected in all healthy blood donor samples in which it occurred with a VAF of > 35%. In addition, variants were excluded if their VAF did not exceed the estimated VAF noise threshold. The VAF noise threshold was calculated individually for each variant as follows: VAFs of variants observed in ≥ two healthy blood donors were ranked from highest to lowest. The highest VAF was discarded and the second highest VAF was multiplied by 1.25, which defined the VAF noise threshold. The 1.25-fold multiplier was chosen empirically to provide a conservative noise threshold, minimizing the risk of falsely excluding low-frequency somatic variants while effectively filtering recurrent background noise observed in healthy donors. Only variants with a VAF above this threshold were retained. Detailed PoN filtering criteria are provided in Supplementary Fig. S1. Variants are reported according to the Human Genome Variation Society (HGVS) nomenclature guidelines based on the GRCh38 reference genome (42).

After PoN filtering, variants were subjected to the following additional tumor assignment workflow to distinguish true tumor-derived mutations from technical artefacts (Fig.1C). Variant calling thresholds were defined according to the availability of prior tumor information. In tumor-informed analyses, previously identified tumor mutations were monitored with a VAF threshold of 0.05% to enable detection of minimal residual disease and early treatment resistance. In the absence of prior tumor information (tumor-uninformed analysis), variants detected by VarDict were required to exceed a VAF threshold of 0.5% to minimize false-positive calls and to comply with the validated lower limit of detection defined by German medical laboratory quality assurance guideline (Rili-BAEK) for routine clinical use (8). In this setting, variants additionally required support by at least three reads and were filtered against the Genome Aggregation Database (gnomAD). If variants occurred in more than 0.1% of the standard population the variant was excluded. No predefined VAF threshold was applied for variant calls generated by Mutect2 (GATK v4.5.0.0).

Variants were annotated for pathogenicity and clinical targetability using OncoKB (v6.1, accessed March 2026) (43,44), COSMIC (v103, accessed March 2026) (45) and ClinVar (National Center for Biotechnology Information; accessed March 2026) (46), with classifications interpreted according to the guidelines of the American College of Medical Genetics and Genomics and the Association for Molecular Pathology (ACMG/AMP) (47).

### 2.7 Statistical analysis

Performance of the panel was assessed by calculating sensitivity, specificity, positive predictive value (PPV), negative predictive value (NPV) and overall accuracy using commercially available reference cfDNA samples (Seraseq ctDNA v2 Mutation Mix, HiSS Diagnostics GmbH, Freiburg, Germany) as the reference standard. In accordance with the manufacturer’s recommendations, variants with a VAF of approximately 50%, consistent with germline variants of the respective cell line, were excluded. True positives (TP) were defined as vendor-documented mutations present in the reference material and correctly detected by the LION panel. False positives (FP) were variants detected by the panel that were not present in the reference cfDNA. However, these variants additionally detected in the vendor’s samples likely represented undocumented true variants rather than technical artefacts. False negatives (FN) were defined as pre-identified mutations not detected by the panel. The total targeted region of the panel comprised ∼500,000 bases. All performance metrics were calculated at a per-base level within the targeted region. True negatives (TN) were calculated as target size - (TP+FP+FN). Analytical sensitivity was calculated as TP/(TP+FN); analytical specificity as TN/(TN+FP); PPV as TP/(TP+FP); NPV as TN/(TN+FN) and accuracy as (TP+TN)/target size. Correlation analyses and Bland-Altman plots (48) were additionally performed to evaluate agreement between NGS-derived VAFs and ddPCR measurements.

## 3 Results

### 3.1. Development of a custom NGS panel, bioinformatics and data analysis workflow (LION panel and workflow)

The LION panel and pipeline were developed as an integrated clinical workflow for liquid biopsy-based molecular profiling in the context of molecular tumor boards (scheme depicted in Figure 1). Briefly, cfDNA is extracted from plasma and subjected to library preparation and hybridization capture-based NGS using the LION panel, a 109-gene targeted NGS panel covering oncologically relevant genes across solid tumor entities (Supplementary Table S1). Beyond SNVs and InDels, the panel detects gene fusions and CNVs. To achieve high sensitivity for fusion detection from liquid biopsy, probes were specifically designed to capture intronic regions of fusion-relevant genes, following a previously published strategy (11). Matched germline DNA from buffy coat was sequenced in parallel, whenever possible, to enable germline-informed variant filtering and discrimination of tumor-specific variants from germline variants. The LION pipeline performs alignment, variant calling, annotation, and multi-layered filtering, including exclusion of germline variants in tumor-informed settings and removal of recurrent background noise using a panel of normal (PoN). Clinically relevant variants are subsequently reviewed and interpreted in the MTB.

### 3.2. Validation cohort and sample composition

To validate the LION panel and the bioinformatic workflow, 85 patient samples, 21 healthy blood donor samples and 17 reference sample replicate measurements were initially screened for inclusion (n=123) (see 2.2.). After applying predefined inclusion and quality control criteria (plasma volume > 1 ml, DNA concentration above the assay detection limit and availability of clinical data), the final validation cohort yielded 87 samples (see CONSORT flowchart, Supplementary Fig. S2, and quality control analysis, Supplementary Fig. S3). The final cohort included 17 reference sample replicate measurements of varying %VAF and DNA input from pre-isolated commercial cfDNA references, 21 plasma-derived cfDNA samples from healthy blood donors and 49 samples from 42 patients. The patient-derived samples consisted of 30 liquid biopsy plasma-derived cfDNA samples, 13 liquid biopsy buffy coat-derived gDNA samples (seven germline-control gDNA samples from cancer patients and six germline-pathogenic gDNA samples from hereditary alpha-tryptasemia (HaT)/mastocytosis patients carrying a germline cKIT mutation), and 6 solid biopsy tissue-derived FFPE samples. The final cohort therefore comprised patient-derived baseline, follow-up, germline gDNA samples and technical replicates as well as healthy blood donor samples and technical replicate measurements of commercial cfDNA reference materials. In total, 28 baseline patient samples were included, comprising 25 baseline plasma samples and 3 baseline tissue FFPE samples. All baseline patient samples had previously been analyzed by WES, ddPCR or other NGS panels. To reflect the intended use of the LION panel in routine MTB diagnostics across a broad spectrum of malignancies, it was validated on this diverse cohort including multiple tumor entities and sample types, as summarized in Table 1.

**Table 1.**
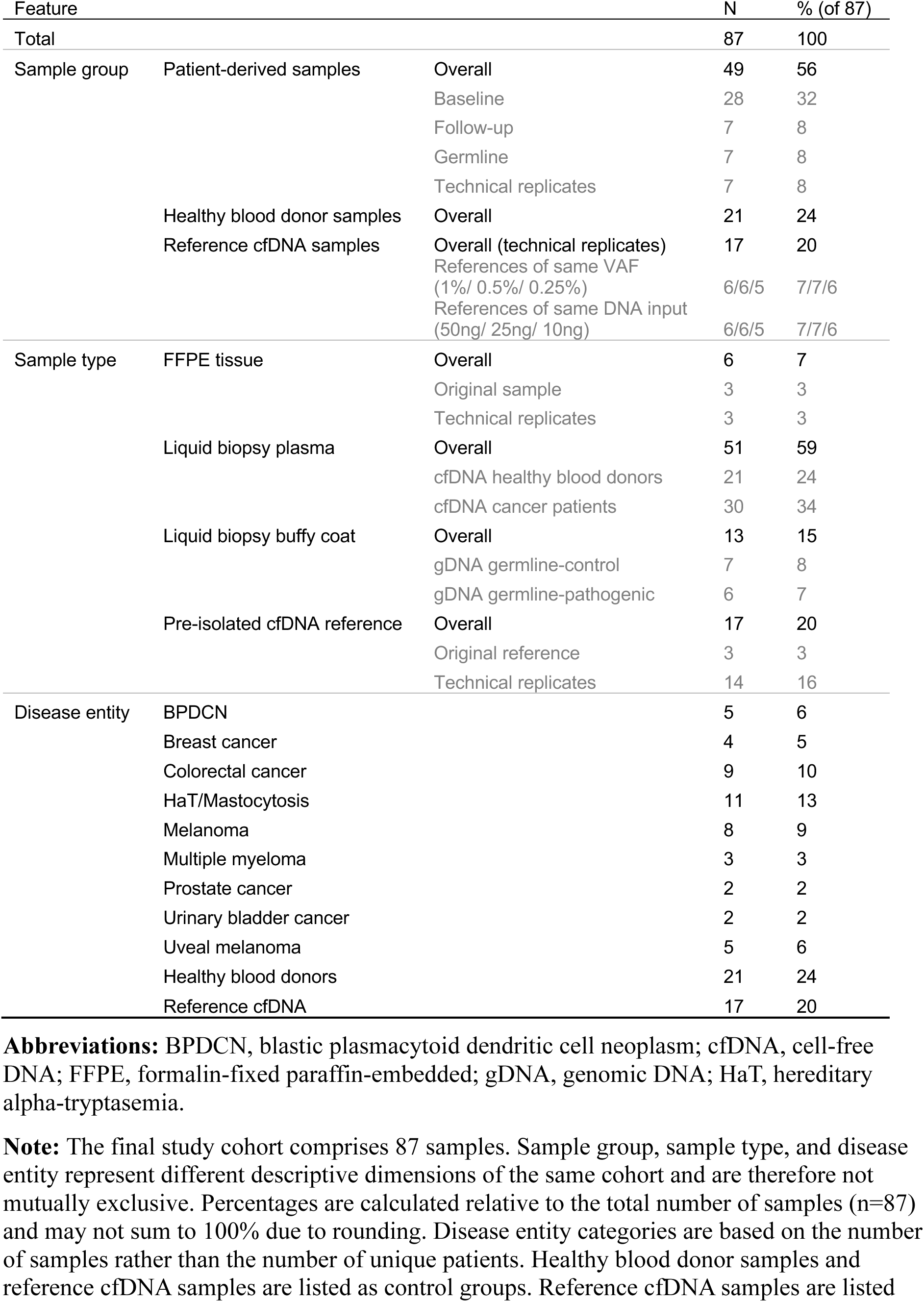
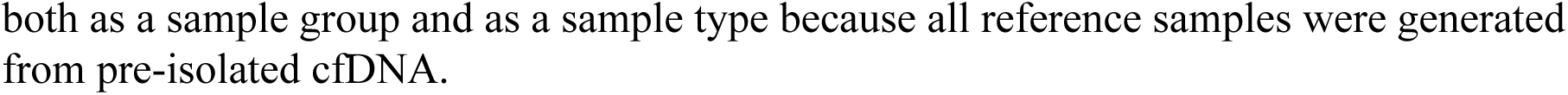
Characteristics of the study cohort.

### 3.3. Sequencing Statistics

To validate the performance of the LION panel for clinical decision making, sequencing depth was evaluated as a key quality control parameter. Sequencing depth was evaluated across different sample types to assess panel performance. After error correction and remapping of clipped reads (EC+RM), the median on-target sequencing depth across all samples was 1967x (mean: 2034x). A wide range of sequencing depths was observed between sample types under identical sequencing conditions. FFPE samples showed a substantially lower sequencing depth with a median depth after EC+RM of 401x (mean 470x) compared to all other sample types. Among liquid biopsy samples, healthy blood donor samples exhibited the lowest sequencing depth (median 888x, EC+RM), whereas cfDNA samples from patients achieved nearly twice the depth, reaching a median on-target depth of approximately 1600x (EC+RM). The gDNA buffy coat patient samples and cfDNA reference samples demonstrated the highest sequencing depths, with median on-target depths up to 3400x (EC+RM). Detailed sequencing depth metrics for all sample types are provided in Table 2. Individual sample-level sequencing depths are listed in Supplementary Table S2 and a summary across runs is shown in Supplementary Table S3. Despite reduced depth in FFPE samples and differences in sequencing depth across sample types, sequencing coverage remained sufficient for reliable variant detection.

**Table 2.** Summary of sequencing depth metrics across sample types and processing steps.

| Sample Type | Subgroup | Processing Step | Mean On-Target Depth<br>± SD [x] | Median On-Target Depth<br>(IQR) [x] | n<br>(Samples) |
| --- | --- | --- | --- | --- | --- |
| FFPE |  | Raw Reads | 4723 ± 887 | 3611 (344) | 6 |
|  |  | EC+RM | 470 ± 264 | 401 (43) | 6 |
| Liquid Biopsy | Overall | Raw Reads | 28005 ± 5940 | 25921 (7668) | 81 |
|  |  | EC+RM | 2150 ± 1357 | 2027 (2062) | 81 |
|  | gDNA Patient | Raw Reads | 28019 ± 4571 | 25921 (5676) | 13 |
|  |  | EC+RM | 3798 ± 1009 | 3408 (1001) | 13 |
|  | cfDNA Patient | Raw Reads | 29294 ± 5834 | 27189 (7396) | 30 |
|  |  | EC+RM | 1680 ± 897 | 1665 (1282) | 30 |
|  | cfDNA Reference | Raw Reads | 26682 ± 4548 | 23946 (6379) | 17 |
|  |  | EC+RM | 3172 ± 927 | 2991 (1153) | 17 |
|  | cfDNA Healthy | Raw Reads | 27227 ± 7621 | 25723 (6598) | 21 |
|  |  | EC+RM | 972 ± 680 | 888 (1320) | 21 |
| **Overall (All Samples)** |  | **Raw Reads** | **26399 ± 8251** | **24913 (7636)** | **87** |
| **Overall (All Samples)** |  | **EC + RM** | **2034 ± 1379** | **1967 (2178)** | **87** |

**Abbreviations:** cfDNA, cell-free DNA; EC+RM, reads after error correction (including deduplication) and remapping of clipped reads; gDNA, genomic DNA; IQR, interquartile range; Raw Reads, reads after mapping but before deduplication; SD, standard deviation.

### 3.4. Concordance of mutations in clinical samples with ddPCR

The mutational detection performance of the panel was evaluated with real clinical samples to assess whether it reliably detects variants with a low false-positive rate and whether the sequencing depth was adequate for identifying low VAFs. The first step was to determine whether the LION panel delivered results comparable to the diagnostically well-established ddPCR method measured from the same cfDNA isolate. To this end, 32 mutations across 28 liquid biopsy samples were compared between the two methods using two different variant callers Mutect2 and VarDict (Fig. 2A-B). The overall sensitivity of the LION panel was 77% for tumor-uninformed analysis and 83% for tumor-informed analysis, with respect to concordance with ddPCR in clinical samples. All mutations with a VAF >0.5% as measured by ddPCR (n=21) were also detected by the LION panel, corresponding to a sensitivity of 100%. In contrast, the sensitivity for mutations with a VAF ≤0.5% (n=9) in tumor-informed analysis was 44%, indicating reduced detection performance at low VAFs. Notably, two *BRAF V600E* mutations were not detected in plasma cfDNA samples by either the LION panel or ddPCR. Correlation analysis demonstrated a strong agreement between NGS and ddPCR measurements from the same sample and same cfDNA isolate, with an r of 0.99 for both variant callers (VarDict and Mutect2). To assess not only correlation but also potential systematic differences between NGS and ddPCR measurements, a Bland-Altman analysis was performed (Fig. 2C-D). The analysis revealed a mean bias (average difference between methods) of 0.25 percentage points for VarDict and 0.18 percentage points for Mutect2 variant caller. The 95% limits of agreement (range containing 95% of the differences) spanned from -3.1 to 3.6 (VarDict) and -3.1 to 3.4 (Mutect2) percentage points. Overall, both variant callers showed comparable agreement with ddPCR.

**Figure 2.**
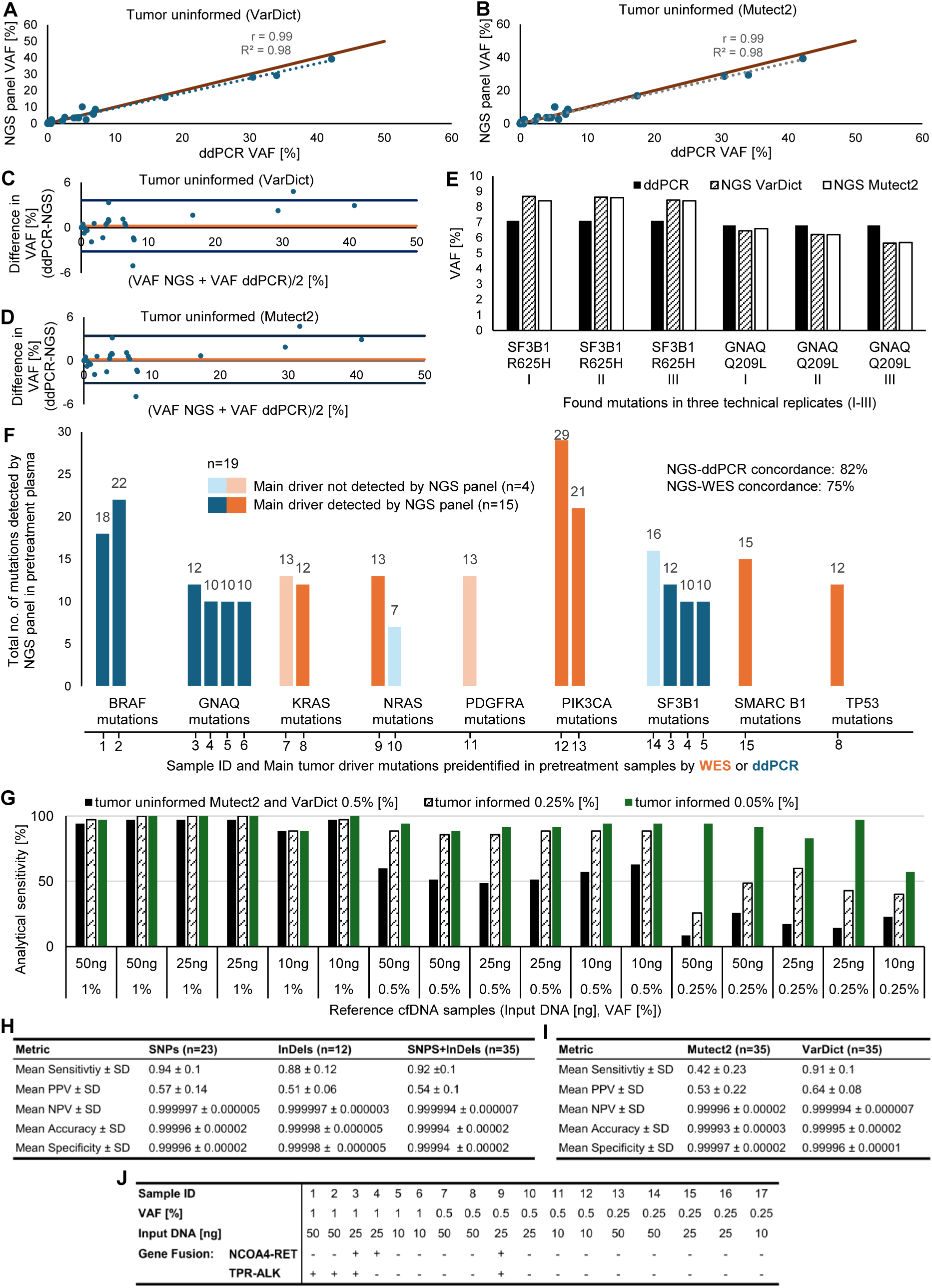
Analytical Performance of the LION NGS panel. (A-B) Correlation of variant allele frequencies (VAFs) measured by ddPCR and NGS for 32 mutations across 28 liquid biopsy samples using VarDict (A) and Mutect2 (B) filters. The brown line indicates the bisector, and the grey dashed line represents the linear regression trendline. (C-D) Bland-Altman plots comparing VAFs measured by the NGS panel and ddPCR using VarDict (C) andMutect2 (D). The orange lines indicate the mean difference (bias; NGS-ddPCR), and blue lines represent the 95% limits of agreement (±1.96 SD). (E) Comparison of VAFs detected by ddPCR and NGS for two mutations across three technical replicates (I-III) of a uveal melanoma patient sample. (F) Distribution of tumor driver mutations detected by the NGS panel in pretreatment samples previously identified by WES (orange) or ddPCR (blue). The NGS panel detected 15 of 19 known main driver mutations (dark bars). Bars represent the number of additional mutations identified by the NGS panel per sample in the same or other genes. (G) Analytical sensitivity of reference cfDNA samples. 35 of 40 pre-identified mutations were analyzed (two fusions evaluated separately; three mutations not covered by the panel). Mutations per sample had VAFs of 1%, 0.5% or 0.25%, with DNA input ranging from 50 to 10 ng. For each sample, three bars represent different filtering criteria. (H) Performance metrics of the NGS panel for SNP and InDel detection. (I) Comparison of performance metrics between Mutect2 und VarDict filter for SNP and InDel detection. (J) Detection of two known gene fusions in reference cfDNA samples. Break support ranged from 4-9 reads for ALK-TPR and from 8-26 reads for NCOA4-RET. “+” indicates detected fusions; “-” indicates fusions not detected. All samples were anonymized and numbered consecutively. ddPCR, digital droplet polymerase chain reaction; ID, identification number; InDel, insertion/deletion; ng, nanogram; NGS, next generation sequencing; No., Number; NPV, negative predictive value; PPV, positive predictive value; SD, standard deviation; SNP, single nucleotide polymorphism; VAF, variant allele frequency.

To assess the reproducibility of our LION panel, we analyzed technical replicates that were prepared either by performing the library preparation on two aliquots of the same sample on the same day or by sequencing two aliquots of the same library preparation in the same sequencing run or in consecutive sequencing runs on different days. High reproducibility was demonstrated across technical replicates prepared and measured on the same day or on different days. As an example, the VAFs of the *SF3B1 R625H* and *GNAQ Q209L* mutations in a uveal melanoma sample, previously detected by ddPCR, were consistently reproduced across replicates (Fig. 2E). For *SF3B1 R625H*, the standard deviation across the three technical replicates was 0.12% for both VarDict and Mutect2 variant callers. For *GNAQ Q209L*, the standard deviation was 0.41% using VarDict and 0.45% using Mutect2. The mean difference between VAF measurements by NGS and ddPCR results was 0.83% for *SF3B1 R625H* and 0.39% for *GNAQ Q209L*. These results demonstrate that the LION panel produces highly reproducible VAF measurements, consistent with those obtained by ddPCR.

### 3.5. Reference and clinical concordance of the LION panel

To evaluate the reliability of mutations detected by the LION panel, its performance specifically at low VAFs (1%-0.25%) was assessed, reflecting conditions observed in early disease settings (Fig. 2G-I). Performance metrics were determined using a commercially available reference cfDNA (Seraseq ctDNA v2 Mutation Mix, HiSS Diagnostics GmbH, Freiburg, Germany). Overall, the panel achieved a mean sensitivity of 92% and a mean specificity of 99% across all conditions. A total of 17 replicates of three reference cfDNA samples containing variants at VAFs of 1% (six replicates), 0.5% (six replicates) and 0.25% (five replicates) were analyzed using input DNA amounts of 50 ng, 25 ng and 10 ng. Technical replicates were measured for all conditions, except for the 0.25% VAF at 10 ng input DNA, for which only a single measurement was obtained due to insufficient sequencing output. The panel detected 35 of 40 mutations present in the reference cfDNA. Three of the five undetected mutations were not covered by the panel, while the remaining two were fusions that were excluded from the performance evaluation and analyzed separately. Sensitivity depended on VAF, DNA input amount and the applied variant filter. Using tumor-uninformed variant calling, sensitivity decreased markedly at VAFs of 0.5% and below, consistent with the applied detection threshold of 0.5% in the VarDict variant caller. Mutect2 showed a lower overall sensitivity compared with VarDict (Fig. 2I) but achieved higher detection rates at low VAFs when compared to tumor-uninformed VarDict analysis (see Supplementary Fig. S4A). Tumor-informed analysis resulted in sensitivity exceeding 90% for most samples and above 82% for all conditions except the 0.25% VAF with 10 ng input DNA, which showed a sensitivity of 57.2%, indicating a lower detection limit. No consistent pattern was observed among false-negative mutations, suggesting the absence of a systematic error (see Supplementary Fig. S4B). Specificity (mean 99%), accuracy (mean 99%), and NPV (mean 99%) remained high across all DNA input amounts and VAF levels, whereas the PPV was moderate, with a mean of 54%. The moderate PPV likely reflects incomplete variant annotation in the reference material, whereby some variants detected by the panel may represent true but unreported mutations. Additional details on false positives and specificity are provided in Supplementary Fig. S4C and D. The performance metrics for each sample and variant caller are summarized in Supplementary Tables S4 and S5. Collectively, these results demonstrate reliable detection of mutations at low VAFs (1%-0.25%), with high overall sensitivity (92%) and specificity (99%). However, positive LION panel test results below a VAF of 0.5% may warrant further confirmatory follow-up diagnostics or progress monitoring.

Based on the established analytical performance of the panel, we next evaluated its suitability for clinical decision-making. Specifically, we assessed whether the LION panel could detect known main driver mutations in pretreatment plasma patient samples that had been identified either with ddPCR in matched liquid biopsy samples or by WES in tumor tissue from the same patient. In total, 19 known driver mutations across 15 pretreatment samples were analyzed. The LION panel detected 15 of 19 driver mutations, corresponding to a concordance of 82% with ddPCR (9 of 11 mutations) and 75% with WES (6 of 8 mutations). Notably, WES had been performed on tissue samples obtained at different time points, making complete concordance unlikely. Importantly, beyond the known driver mutations, the panel identified an average of 14 additional variants per sample (Fig. 2F), underscoring the added value of the panel for comprehensive molecular profiling in a clinically relevant context.

The LION panel was not specifically designed for fusion gene analysis but the presence of probes spanning the well-described break points, including *ALK*, *RET* and *ROS* as described by Seki et al, enabled fusion detection from liquid biopsy (24). Using reference cfDNA samples, two gene fusions (*TPR-ALK*; *NCOA4-RET*) were known to be present per sample. Fusions were detected by the LION panel at low VAFs (1%-0.25%) in 5 of 17 reference cfDNA samples, with detection rates increasing with higher VAFs and DNA input amounts (Fig. 2J). Breakpoint support for detected fusions ranged from 4-9 reads for *TPR-ALK* and from 11-26 reads for *NCOA4-RET*. Other gene fusions found in reference cfDNA samples showed breakpoint support of up to 484x. All identified gene fusions with break support values exceeding five reads at both breakpoints are listed in Supplementary Table S6.

### 3.6. Mutational landscape of the study cohort

After assessing the analytical and clinical performance of the panel, the mutational landscape of the cohort was analyzed to identify recurrently mutated genes, entity-specific patterns, and potential systematic effects. To this end, a tumor-uninformed analysis of all 87 samples across the 109 genes included in the LION panel was performed. For each sample, the presence of one or more variants per gene was determined and visualized in an oncoplot (Fig. 3; Supplementary Fig. S5). *KMT2D* was the most frequently mutated gene, with variants detected in more than 60% of cohort samples. Notably, variants in this gene were also identified in 14 of 21 healthy blood donor cfDNA samples and in 15 of 17 reference cfDNA samples. *DNMT3A* was the second most frequently mutated gene. Both genes are well-established CHIP-associated genes, consistent with their detection in healthy blood donor samples. As expected, FFPE samples exhibit multiple variants across nearly all analyzed genes, likely reflecting a high prevalence of unspecific sequencing artifacts caused by formalin-induced cytosine-deamination and crosslinking. However, due to insufficient sequencing depth, as described above, FFPE samples were therefore excluded from further analyses. Reference cfDNA samples displayed a mutation pattern consistent with the predefined variants they were designed to contain. However, several genes not specified by the manufacturer as mutated (including *KMT2D*, *DNMT3A* and *TSC1*) were nevertheless identified as mutated by our LION panel in most reference samples. Following inquiry with the manufacturer, it was confirmed that the presence of additional mutations beyond those specified in the reference material could not be excluded.

**Figure 3.**
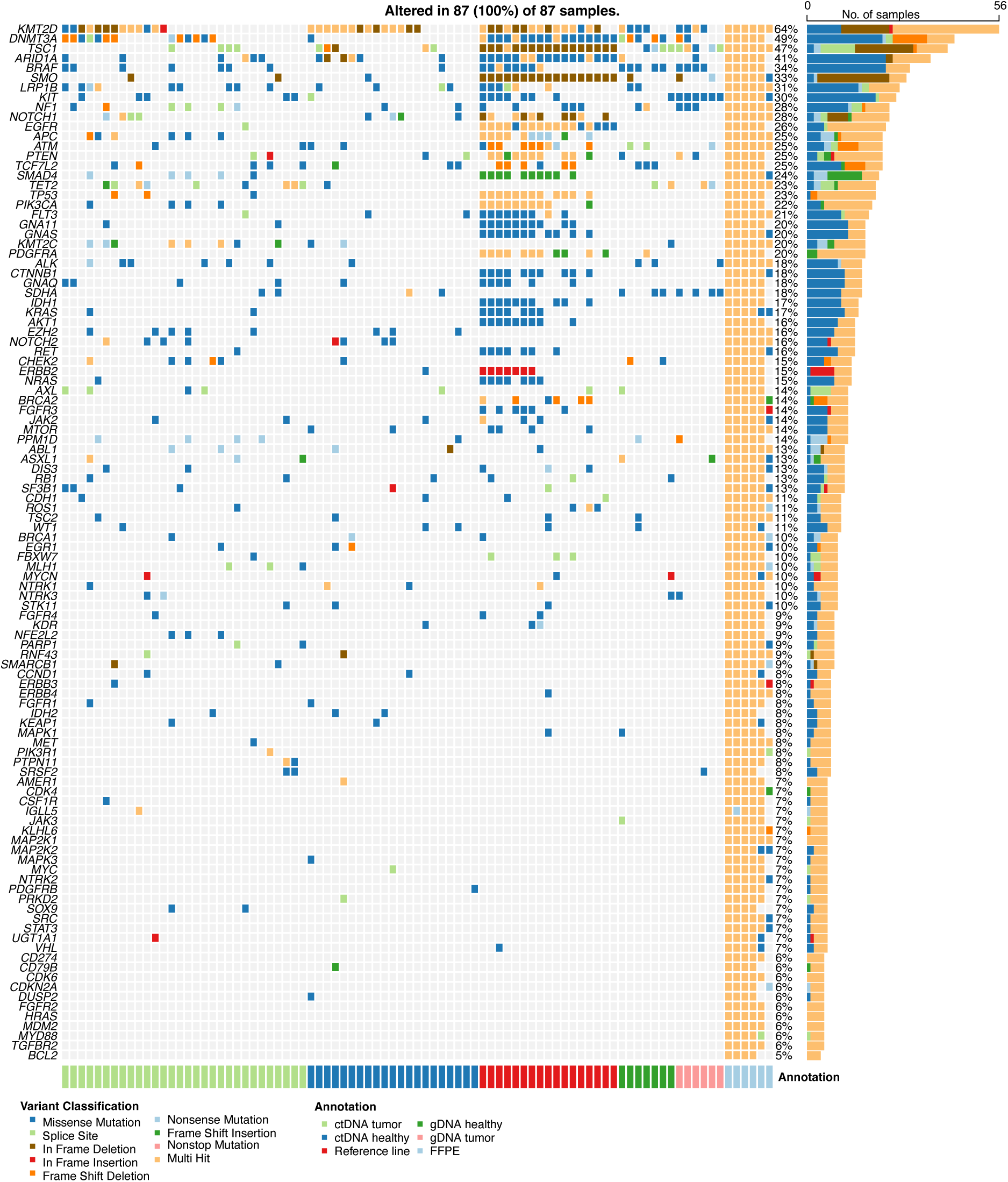
Mutational landscape of analyzed cohort using a tumor-uninformed approach. Rows represent genes and columns represent individual samples. Colors indicate mutation and sample types. ctDNA, circulating tumor DNA; FFPE, formalin-fixed paraffin-embedded; gDNA, genomic DNA.

### 3.7. Longitudinal case studies demonstrate the clinical utility of the LION panel

To illustrate the potential clinical relevance of the LION panel, we describe here three representative cases for which longitudinal clinical follow-up data were available, allowing correlation of disease progression with LION panel sequencing results.

#### 3.7.1. Breast cancer patient case with potentially clinically relevant molecular findings added by LION panel analysis

The first case describes a patient with an estrogen receptor (ER)-positive, progesterone receptor (PR)-positive breast cancer of no special type (NST). At initial diagnosis, breast-conserving surgery and radiation therapy were performed, followed by tamoxifen treatment for three and a half years until the occurrence of the first bone metastases (defined here as month 0). Subsequently, metastasectomy, adjuvant chemotherapy with epirubicin and cyclophosphamide and radiation therapy were carried out. However, over the following four years, two confirmed and two presumed disease progressions necessitated four changes in systemic therapy, including palbociclib, exemestane, alpelisib and paclitaxel (Supplementary Table S7).

At the time of discussion at the MTB, over four years after the initial diagnosis of metastatic disease and following at least the third disease progression, metastases were present in bone and liver. To facilitate targeted therapy, repeated attempts were made to obtain a tissue biopsy. However, two biopsies were unsuccessful due to insufficient material collection. In parallel, at 4 years and 5 months after the first metastasis, a liquid biopsy was collected and analyzed by ddPCR for the *PIK3CA E545K* mutation (Fig. 4A and B). However, detection of the *PIK3CA* variant did not result in additional therapeutic options because the patient had previously progressed on alpelisib treatment during the second year after first metastasis. Following unsuccessful tissue sampling, treatment with capecitabine and denosumab was initiated, resulting in a mixed response. A further attempt to obtain a tissue biopsy from liver metastasis was made, but the DNA quality was inadequate for analysis. To identify further therapeutic options, a WES was performed on a tissue biopsy obtained at the time of the first tumor progression more than five years earlier. Based on these results, treatment with trastuzumab deruxtecan was administered, followed by sacituzumab govitecan. Despite these interventions, the patient experienced further disease progression and died approximately six years after the first metastatic progression. In contrast to tissue biopsies, liquid biopsies could be collected reliably and ddPCR successfully detected the tumor-derived *PIK3CA E545K* variant. However, unlike NGS analysis, ddPCR was limited to this single mutation and therefore did not reveal additional therapeutic options, particularly as alpelisib had already been administered.

**Figure 4.**
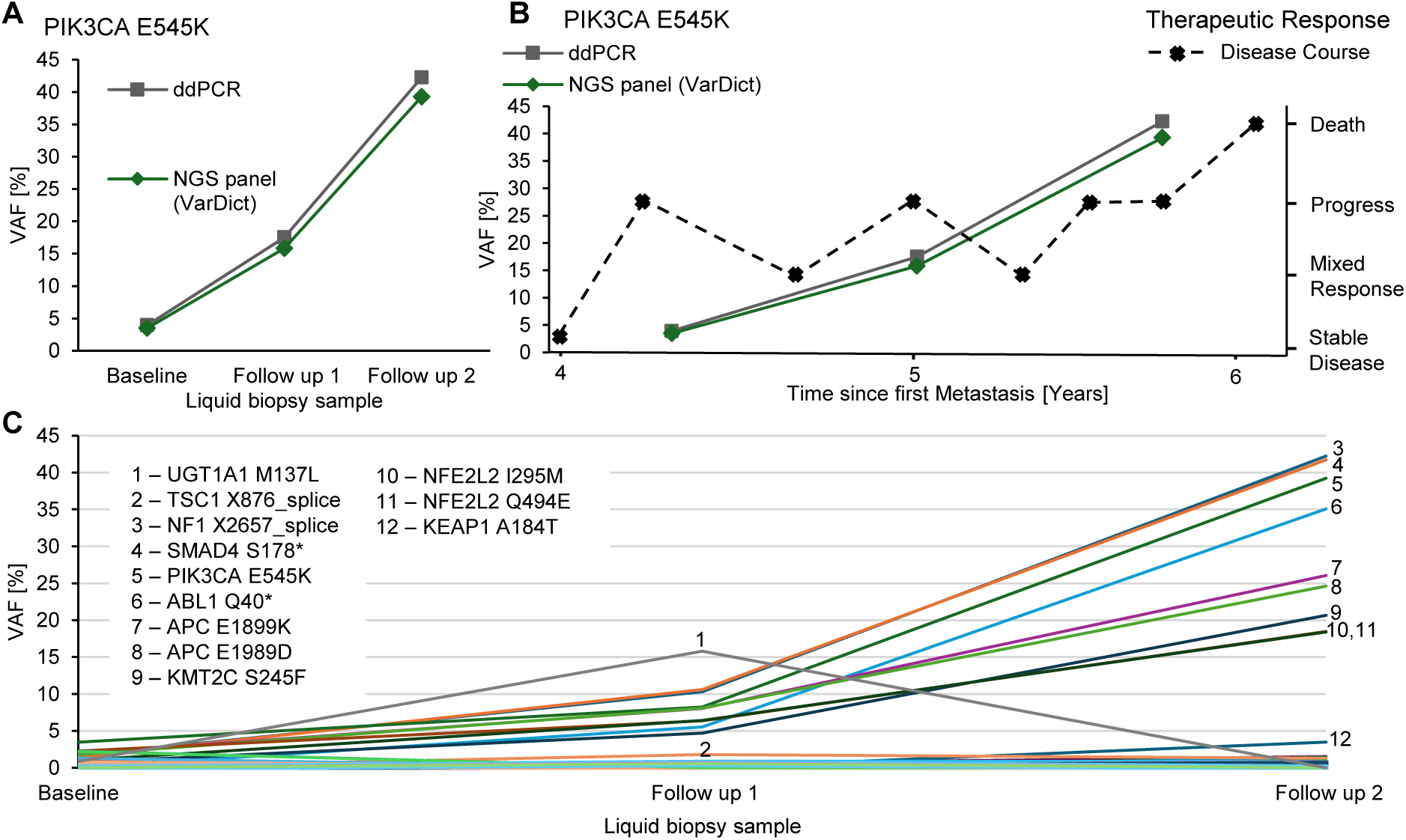
Mutational profile, disease course and therapeutic response in liquid biopsy (cfDNA) samples from a breast cancer patient. (A) Longitudinal comparison of PIK3CA E545K detection in serial breast cancer patient samples by ddPCR and NGS (VarDict filter). (B) Dynamics of PIK3CA E545K VAF in relation to therapeutic response. Clinical response data were taken directly from the physician report (Supplementary Table S7). Lines connecting points are shown for visual guidance only and do not represent continuous or quantitative interpolation. (C) Longitudinal mutational landscape detected by the LION panel in samples of the breast cancer patient. Only mutations with a VAF <45% across all samples are shown. The 12 mutations with the highest VAF (>1.5%) are labeled (Supplementary Figure S6 and Table S8A). cfDNA, cell-free DNA; ddPCR, digital droplet polymerase chain reaction; NGS, next generation sequencing; VAF, variant allele frequency.

The next step was to evaluate whether the LION panel could reliably detect the previously identified *PIK3CA E545K* variant and could reflect disease dynamics and identify additional variants of potential therapeutic relevance. LION panel analysis showed high concordance with ddPCR for detection of the *PIK3CA E545K* variant (Fig. 4A). The ddPCR revealed that the VAF increased over time and correlated with therapeutic response (Fig. 4B). In addition to *PIK3CA E545K*, the LION panel detected 32 further variants in plasma samples of this patient. Nine of these variants showed VAF dynamics concordant with *PIK3CA E545K* (Fig. 4C), suggesting tumor association and potential biological relevance. Detailed information on all detected variants with a VAF <45% in cfDNA samples (somatic variants) is provided in Supplementary Fig. S6. To assess the contribution of potential germline and CHIP variants, an additional analysis of gDNA of this patient was performed. If a patient is CHIP positive, identical mutations (not necessarily at identical VAFs) are expected to be present in both gDNA derived from peripheral blood leukocytes and cfDNA. In contrast, mutations in CHIP associated genes or potential germline variants detected exclusively in cfDNA are more likely to represent tumor-derived variants independent of CHIP or germline background, for example mutations involved in carcinogenesis (e.g. TP53 mutations). The gDNA sample was isolated from the follow-up 2 sample. Variants in *NOTCH2* and a *CHEK2* variant were detected in the gDNA and also in all cfDNA samples with a VAF >45%, which is compatible with a germline origin (Supplementary Table S8A). Six additional variants were detected in the gDNA with a VAF <1%, four of which (*TSC1 X876_splice*, *BRAF E26A*, *DNMT3A M761V* and *PIK3CA E545K*) were also detected in a subset of cfDNA samples, consistent with CHIP-associated mutations. The *PIK3CA E545K* variant was detected in the gDNA sample with a VAF of 0.09%, which is likely attributable to a plasma cfDNA contamination, given the high VAF observed in the corresponding cfDNA follow-up 2 sample. Furthermore, we also analyzed the fusion gene results of this patient (Supplementary Table S8B). We found one fusion (*ANKRD10-PARP1*) exceeding five reads at both breakpoints in the two follow-up samples. In conclusion, the LION panel enabled the detection of multiple tumor-associated variants beyond standard tissue-based genotyping, thereby providing a more comprehensive molecular profile in a clinically challenging breast cancer case with repeated failure of tissue sampling. In the Discussion section, potential therapeutical implications and options for this molecular profile will be reviewed.

#### 3.7.2. Melanoma cancer patient case with potentially clinically relevant molecular findings added by LION panel analysis

The second case describes a patient with cutaneous melanoma (Fig. 5A-C and Supplementary Fig. S7), illustrating longitudinal cfDNA profiling using a minimally invasive liquid biopsy approach to capture molecular disease evolution over time. A baseline liquid biopsy was obtained at the time of initial diagnosis, followed by three longitudinal samples collected at week 2, 16, and 18 after initial diagnosis. All four cfDNA samples were analyzed by ddPCR for the presence of the recurrent melanoma driver mutation *BRAF V600E*. The LION panel results showed a trend comparable to the ddPCR analysis (Fig. 5A). While ddPCR detected *BRAF V600E* in all four samples, the LION panel identified the mutation in three of the four cfDNA samples. In the baseline sample the VAF was 0.1% by ddPCR and 0.5% by NGS. In contrast, the first follow-up sample exhibited a VAF of only 0.04% by ddPCR, which was below the detection limit of the LION panel (see section 3.4). In the third and fourth sample, both ddPCR and NGS detected the mutation. Importantly, longitudinal *BRAF V600E* VAF dynamics showed clear increase over time, which correlated with clinical disease progression (Fig. 5B). This supports cfDNA based VAF quantification as a clinically relevant surrogate marker for tumor burden and disease monitoring in metastatic melanoma. Whereas ddPCR targeted only *BRAF V600E*, the LION panel identified a total of 44 variants across all four cfDNA samples, revealing substantial tumor molecular heterogeneity that is not captured by single-gene testing. Most variants exhibited VAFs below 1%. However, nine variants with VAFs exceeding 1% were highlighted in Fig. 5C. Four variants (labeled 5 to 7 and 9 in Fig. 5C) were already detectable at the time of first metastasis (follow-up 2 sample) and variants 5 to 9 showed increasing VAFs over time. These variants were therefore selected for further investigation as potentially biologically relevant. In addition, an *ALK V811M* variant was detected in all samples with VAFs ranging from 43% - 48%, suggesting a germline origin. However, a matched gDNA sample was not available for CHIP or germline confirmation. Fusion gene analysis was also performed. We found one fusion (*LINC00486-KMT2D*) exceeding five reads at both breakpoints in follow-up sample 1 and 2. The results are summarized in Supplementary Table S9. In conclusion, longitudinal cfDNA profiling using the LION panel enabled minimally invasive monitoring of molecular tumor evolution across different melanoma disease stages using liquid biopsy. Compared with single time-point analyses, serial sampling more effectively captured dynamic clonal changes and emerging potentially targetable variants during disease progression. The therapeutical implications will be explored in the Discussion section.

**Figure 5.**
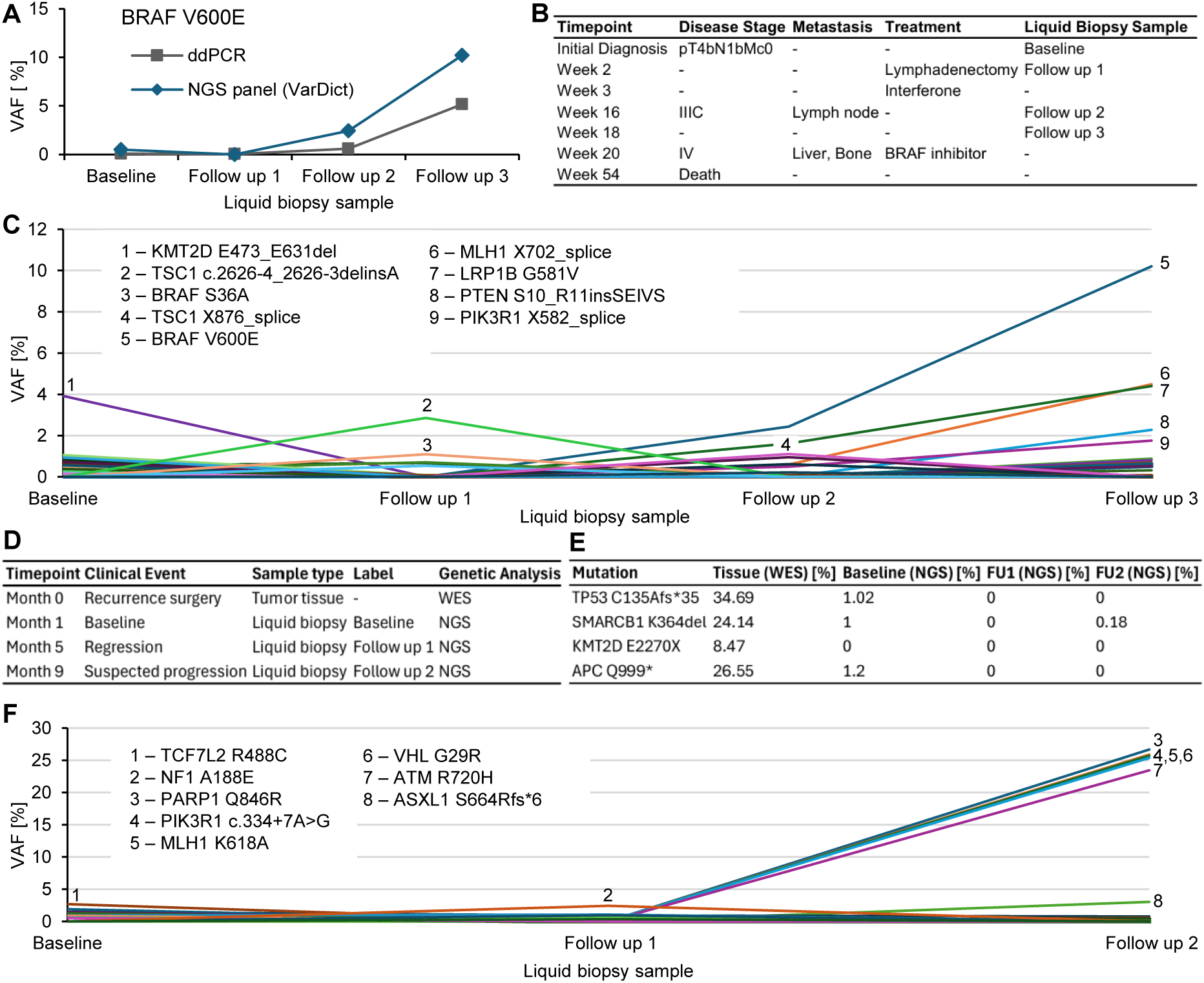
Mutational profile and disease course in a melanoma (A-C) and a rectal cancer patient (D-F). (A) Longitudinal comparison of BRAF V600E detection in liquid biopsy samples from the melanoma patient using ddPCR and NGS (VarDict filter). In NGS analysis, the baseline value represents the average of two technical replicates. (B) Overview of sample collection, treatments and disease stages for the melanoma patient. Time points are indicated as weeks after initial diagnosis. Disease stage was determined clinically according to the AJCC classification (2009). Metastasis indicates the presence and location of metastases. All samples are liquid biopsies (cfDNA). (C) Longitudinal dynamics of mutations detected by the LION panel in liquid biopsy samples from the melanoma patient. The baseline sample consists of two technical replicates. Only mutations with a VAF <40% across all samples are shown. The 9 mutations with the highest VAF (>1%) are labeled. For additional details, see Supplementary Figure S7. (D) Overview of sample collection and genetic analyses for the rectal cancer patient in relation to clinical status. Timepoints indicate months after recurrence. Clinical events were assessed by imaging (MRI/CT): “Regression” = tumor regression observed; “Suspected progression” = possible disease progression based on clinical findings (suspicious presacral and pulmonary lesions, as well as suspicious mediastinal and para-iliac lymph nodes). (E) Mutations identified in tumor tissue by WES that were also detectable by the LION panel. The baseline liquid biopsy sample was collected one month after surgery for tumor recurrence; FU1 and FU2 were collected five and nine months later, respectively (see Figure 5D). (F) Longitudinal dynamics of mutations detected by the LION panel in liquid biopsy samples from the rectal cancer patient. Only mutations with a VAF <30% across all samples are shown. The 8 mutations with the highest VAF (>2%) are labeled. For additional details, see Supplementary Table S11B and Figure S8. cfDNA, cell-free DNA; ddPCR, digitaldroplet polymerase chain reaction; FU, follow-up; NGS, next generation sequencing; TRA, translocation; VAF, variant allele frequency; WES, whole exome sequencing.

#### 3.7.3. Rectal cancer patient case with potentially clinically relevant molecular findings added by LION panel analysis

The third case describes a patient with rectal cancer who developed recurrent disease nearly two years after initial diagnosis and surgical resection (Fig. 5D-F, Supplementary Fig. S8, Table S10 and Table S11). The baseline liquid biopsy sample was collected one month after surgical resection of the recurrence, followed by collection of two longitudinal liquid biopsy samples at month 5 and 9 (Fig. 5D). While ddPCR analysis was not available for this patient, WES of an FFPE tumor tissue sample obtained during surgery of the recurrence identified four mutations covered by the LION panel (*TP53 C135Afs*35*, *SMARCB1 K364del*, *KMT2D E2270X*, *APC Q999\**), although the WES analysis was limited to variants with ≥ 5% VAF and thereby limiting the detection of low VAF mutations. Three of the four mutations were detectable in the baseline liquid biopsy collected one month after surgery despite a low VAF of ≤1.2% (Fig. 5E), indicating the presence of MRD that would likely remain undetected by less sensitive approaches. The patient underwent radiation therapy between month 1 and month 3 after recurrence surgery, which was associated with radiologic regression at month 5. However, longitudinal LION panel analysis revealed the emergence of additional variants over time (Fig. 5F). Variants labeled 3 to 8 showed increasing VAFs in follow-up 2 sample and were therefore considered most consistent with progressive clonal expansion. Importantly, these molecular findings are consistent with clinically suspected disease progression, as presacral and pulmonary lesions as well as mediastinal and para-iliac lymph-nodes were identified as suspicious, ultimately leading to initiation of palliative systemic treatment with FOLFOXIRI and panitumumab. Follow-up imaging four months later confirmed presacral recurrence. To further assess the origin of detected variants, matched gDNA analysis from the baseline liquid biopsy sample was performed. Ten variants were detected in the gDNA sample, nine of which showed VAFs < 2.5%. The *RB1 F293L* variant showed a VAF of approximately 50% across baseline and follow-up 1 samples, which is consistent with a germline origin. However, its VAF decreased to 22% at follow-up 2, which may reflect technical variability due to reduced cfDNA input, or alternatively suggest a somatic origin with clonal dynamics. The variants *DNMT3A W306Cfs*10* and *TET2 X1349_splice* were also detected in cfDNA, indicating potential CHIP. All remaining variants were detected exclusively in gDNA (Supplementary Table S11A). In addition, fusion gene analysis was performed. We found one fusion (*LINC00486-KMT2D*) exceeding five reads at both breakpoints in follow-up sample 1. The results are provided in Supplementary Table S11B. In conclusion, this case illustrates the potential clinical utility of the LION panel for highly sensitive post-surgical ctDNA monitoring in rectal cancer. The analysis enabled detection of residual disease at low VAFs shortly after surgical resection and captured dynamic clonal evolution preceding radiologically confirmed progression, thereby highlighting its value as a complementary approach to tissue-based genotyping for longitudinal disease surveillance and therapeutic decision support.

## 4 Discussion

In this study, we validated a customized pan-cancer liquid biopsy open NGS panel combined with an in-house bioinformatic workflow (LION panel and pipeline) for clinical decision-making specifically designed for use in MTBs. The LION panel reliably detects clinically relevant cancer-associated mutations in cfDNA and gDNA used for targeted molecular testing and germline control purposes, including low VAF variants, and enables sensitive mutational profiling even in samples with limited ctDNA input. Importantly, the LION panel provides clinically actionable insights that directly support MTB decision-making: It enables longitudinal monitoring of MRD, early detection of recurrence, and identification of emerging resistance mutations. Furthermore, it facilitates therapy stratification based on dynamic ctDNA profiles, particularly in patients where tumor tissue biopsies are unavailable or insufficient.

Sequencing depth was optimized to balance sensitivity and cost-effectiveness (∼350€ per sample). Based on a standardization study by Petrackova et al, a minimum depth of 1650x is recommended for a limit of detection of ≥3% VAF, while lower VAFs (1-3%) require confirmation (49). Here in our study, all liquid biopsy patient and reference cfDNA samples exceeded this threshold. Sequencing depth varied depending on sample type and DNA characteristics, with lower median depth (∼888x) in healthy cfDNA after error correction likely due to reduced DNA input and library complexity (50–52). For FFPE samples the median on-target depth was 401x, while showing increased mutation calls, consistent with formalin-induced artefacts (53,54). Although median on-target depth was generally high across liquid biopsy samples, coverage variability (IQR) remained substantial, consistent with known cfDNA fragmentation and low-input effects (55). Importantly, the analytical sensitivity of NGS-based ctDNA analysis is not only determined by sequencing depth but also by the limited number of genome copies present in the cfDNA input. Accordingly, 10ng cfDNA corresponds to a theoretical lower limit of detection of approximately 0.03% VAF, even under ideal conditions of complete library conversion and unlimited sequencing depth (56).

Analytical validation using reference cfDNA demonstrated high sensitivity (92%), specificity (99%), accuracy (99%) and NPV (99%). The comparatively moderate PPV (54%) is likely underestimated due to incomplete variant annotation in the reference material, leading to misclassification of true variants as false positives (57). In addition, missing prevalence data limits assessment of both PPV and NPV (58,59). Fusion detection remained limited, particularly at low VAFs, reflecting known challenges of DNA-based fusion analysis due to heterogeneous breakpoint regions (20,60).

Compared to other comprehensive ctDNA assays of comparable size (∼100 genes), such as commercially available AlphaLiquid100 and FoundationACT, the LION panel demonstrates comparable analytical performance while providing the additional advantage of a fully transparent, manufacturer-independent, and freely available workflow. In contrast to commercial solutions, which often generate large numbers of clinically non-actionable mutations and thereby increase analytical complexity and costs (23), the LION panel is specifically designed for application in real-world MTB workflows, where analytical efficiency, clinical interpretability, and cost-effectiveness must be balanced across diverse patient cohorts. Across studies, ctDNA assays of this size generally achieve high sensitivity and specificity, although reported PPV values vary. The limit of detection typically ranges between 0.1% and 0.5% VAF (61–64), which is consistent with real-world NGS-based ctDNA data in NSCLC (65). For variants near lower detection thresholds, validation using ddPCR has been suggested to improve analytical confidence (66). In this context, the LION panel indicates the potential for improved sensitivity at low VAFs, likely enabled by increased sequencing depth and optimized error correction strategies. The associated bioinformatic workflow applied in this study shares key features with previously published sequencing pipelines, including advanced variant filtering and artefact reduction strategies (67).

A key feature of the LION pipeline is the parallel use of two complementary variant callers, VarDict and Mutect2, which differ in their sensitivity and specificity profiles at low VAFs. While VarDict demonstrated higher overall sensitivity, Mutect2 provided complementary detection at very low VAFs in tumor-uninformed settings (Fig. 2I; (68)). UMI-based error correction (69) was implemented to reduce PCR and sequencing errors, which is essential for reliable detection of variants at VAFs approaching the noise floor. A panel of normals derived from 21 healthy blood donors was incorporated to filter recurrent background variants, providing a sample-type-specific noise model that complements standard population databases such as gnomAD. Together, these features enable the LION pipeline to operate effectively across a range of clinical scenarios, from tumor-uninformed screening to highly sensitive tumor-informed MRD monitoring.

Although regulatory guidelines currently mandate validation of NGS panels down to 0.5% VAF (8), clinically relevant variants may occur below this threshold (70,71) and may therefore escape detection in guideline-compliant workflows. In this study, concordance between the LION panel and ddPCR was high (r = 0.99), although sensitivity decreased at 0.5% VAF, consistent with previous reports (72). For predefined variants, the panel achieved improved sensitivity, detecting variants down to 0.05% VAF and reaching 97% sensitivity at 0.25% VAF, depending on DNA input. This level of sensitivity enables monitoring of MRD and early detection of treatment resistance, since positive ctDNA after treatment predicts disease relapse (14).

Bland-Altman analysis (48) indicated minimal systematic bias between NGS and ddPCR measurements with moderate variability (95% limits of agreement; -3.1 to 3.6 percentage points) consistent with previous reports comparing NGS and ddPCR (66,73). This variability is likely negligible at higher VAFs but may affect measurements at lower VAF levels. Notably, variants previously detected by ddPCR were identified with a 100% sensitivity at VAFs >0.5%, further supporting the suitability of the LION panel for detecting low-VAF variants.

Pretreatment plasma analysis showed high concordance with ddPCR (82%) and WES (75%) for driver mutations, consistent with, and in part exceeding, previously reported agreement between NGS and established methods (66,74,75). WES had been performed on tissue samples obtained at different time points, making complete concordance unlikely. Discordant cases were primarily attributable to low VAFs (<0.5%) or temporal and spatial tumor heterogeneity. Importantly, the LION panel identified additional variants (mean 14 per sample), thereby expanding the molecular landscape beyond predefined targets, supporting the clinical utility of the LION panel for liquid biopsy-based mutational profiling.

Frequent mutations in *KMT2D* and *DNMT3A* were observed across all 87 samples of the cohort, including healthy controls and reference material. These genes, particularly *DNMT3A*, are known to be associated with CHIP (76–78). Differentiation between CHIP- and tumor-derived variants is essential in tumor-uninformed analyses. To address this, we accounted for CHIP-associated genes included in our panel and, whenever possible, used matched leukocyte-derived gDNA patient samples as control. A CHIP mutation is expected to be detected in both cfDNA and matched leukocyte-derived gDNA. If gDNA is missing, longitudinal monitoring of CHIP-associated variants using the LION panel may further aid in their interpretation. In contrast to CHIP-associated mutations, tumor-derived variants are expected to show dynamic changes in VAF that correlate with disease progression or treatment response (79,80). Failure to account for CHIP may lead to false positive MRD detection and could impact clinical interpretation.

Longitudinal monitoring of patients with advanced, pretreated cancer was feasible in all three case studies, primarily due to the minimally invasive and repeatable nature of liquid biopsy compared to tissue biopsy. Liquid biopsy is particularly valuable in settings where tissue collection is unsuccessful, or imaging findings are inconclusive. VAF dynamics measured by the LION panel correlated with disease progression and treatment response. Dynamics of ctDNA have been shown to correlate with tumor burden, with decreasing levels indicating response and increasing levels suggesting progression or recurrence (14,81–83). While ddPCR enables highly sensitive detection of predefined variants, the LION panel allows comprehensive monitoring of tumor evolution and emerging mutations. In contrast, tissue-based WES provides only a static snapshot, which may underestimate disease progression and miss newly emerging clinically actionable variants.

Across all cases, variants were systematically classified as likely somatic tumor-derived, germline, or CHIP-associated. Germline variants were defined by their presence in gDNA and/or consistently high VAF (>30%) in cfDNA across longitudinal samples without tumor-associated dynamics (84). Importantly, such variants may still be clinically relevant for therapeutic decision-making.

Potentially targetable variants were identified in all case studies based on mutational dynamics. In the breast cancer patient, *PIK3CA E545K* was already therapeutically exhausted, but nine additional variants detected by the LION panel showed dynamic changes suggesting alternative targets: *NF1 X2657_splice*; *SMAD4 S178\**; *ABL1 Q40\**; *APC E1899K*; *APC E1989D*; *KMT2C S245F*; *NFE2L2 I295M*; *NFE2L2 Q494E* and *KEAP1 A184T*. *NF1 X2657_splice*, is annotated in OncoKB (43,44) as a likely oncogenic loss-of-function somatic mutation and has previously been reported in breast cancer (85). Notably, this variant showed the highest VAF (42.2%) in the final follow-up sample, indicating a strong association with tumor progression. This variant may qualify for off-label treatment with MEK1/2 inhibitors such as cobimetinib or trametinib, although MEK inhibition is not standard treatment in breast cancer (OncoKB level 4). *KMT2C S245F* may have potential therapeutic relevance, as *KMT2C* deficiency has been associated with increased sensitivity to PARP inhibition (86). In addition, menin inhibitors, which are Food and Drug Administration (FDA) approved for *KMT2A*-rearranged acute myeloid leukemia based on the phase I/II AUGMENT-101 study (NCT04065399), are being explored in other *KMT2* malignancies, although their relevance for *KMT2C* mutations remains to be established (87,88). For the remaining variants, no targeted treatment options are currently available. However, *SMAD4 S178\** also showed tumor-associated dynamics. Although no targeted therapy is currently available, *SMAD4* is an active area of research in metastatic breast cancer and may become therapeutically relevant in the future (89). The subclonal *TSC1 X876_splice* variant (<2% VAF), provides a potential rationale for mTOR inhibition as *TSC1* alterations are treated with everolimus (mTOR inhibitor) as standard of care in glioma (OncoKB level 1). Although of limited relevance at the observed allele frequency, such variants may warrant longitudinal monitoring. Notably, *KEAP1* mutations are often detected in NSCLC and are associated with therapeutic resistance (90). Two germline variants (*NOTCH2 c.751+8G>A*, *CHEK2 R474H*) were identified, neither of which currently offer treatment options.

In the melanoma patient, five variants showed dynamics consistent with disease stage and treatment response, suggesting potentially targetable variants: *BRAF V600E*; *MLH1 X702_splice*; *LRP1B G581V*; *PTEN S10_R11insSEIVS*, and *PIK3R1 X582_splice*. *BRAF V600E* appears to be the dominant driver mutation, with the highest VAF (10.21%) in the final follow-up sample and a marked increase over time, consistent with disease progression. It is classified as an oncogenic gain-of-function mutation, and BRAF inhibition represents standard of care in melanoma (OncoKB level 1). Current standard-of-care guidelines recommend immune checkpoint inhibition (ICI) or alternatively BRAF-and-MEK inhibition (91). Given that the patient’s case dates back to 2010-2011, treatment reflected the standard therapeutic options available at that time, and the patient received interferon-based therapy in week three after diagnosis. A BRAF inhibitor was subsequently administered within the framework of a clinical study at week 20 after initial diagnosis in stage IV melanoma. However, the patient deceased at week 54. The *MLH1 X702_splice* variant had the second highest VAF (4.49%) at the final follow-up sample. OncoKB classifies it as a likely oncogenic loss-of-function mutation. While no single direct targeted therapy exists matching this mutation profile, rapid liquid biopsy examination would have identified the BRAF mutation and the high number of tumor mutations (n=44), and could have allowed standard-of-care guideline based treatment with immune checkpoint inhibitors (ICI) (92). The *PIK3R1 X582_splice* variant is annotated as a likely oncogenic loss-of-function mutation in OncoKB. Preclinical evidence suggests that combined MEK and PIK3CA inhibition may also be effective, supporting a possible therapy rationale (93,94). The remaining two variants, *LRP1B G581V* and *PTEN S10_R11insSEIVS*, currently lack established targeted treatment options. PTEN alterations may also provide a biological rationale for AKT pathway inhibition, although the pathogenicity of this specific variant remains unclear (95). A subclonal *TSC1 X876_splice* variant was detected in follow-up sample 2 with a VAF of 1.11 %, suggesting a minor role in tumor progression. Nonetheless, it is a likely oncogenic loss-of-function mutation, which warrants longitudinal monitoring due to a potential rationale for mTOR inhibition. Although no gDNA was available, the *ALK V811M* variant was present in all longitudinal samples with a VAF > 43%, suggesting a germline origin. ClinVar classifies it as a variant of uncertain significance, and its clinical significance remains unclear.

In the rectal cancer patient, the VAFs of six variants increased at the time point of suspected progression, suggesting potential therapeutic relevance: *PARP1 Q846R*; *PIK3R1 c.334+7A>G*; *MLH1 K618A*; *VHL G29R*; *ATM R720H*; *ASXL1 S664Rfs*6*. Notably, canonical colorectal cancer (CRC) driver genes such as *TP53* and *APC* were only detected in tissue biopsy at the time of recurrence surgery and were not identified in the liquid biopsy LION analysis at the time of suspected progression. Three out of the six potential targetable variants with increasing VAFs are described as recurrently mutated driver genes in CRC: *ASXL*, *PIK3R1* and *MLH1* (96), but no established targeted therapies are currently available. Alterations in *MLH1* may be associated with MSI, which can predict response to ICI. However, MSI status could not be assessed using the LION panel, and the *MLH1 K618A* variant is classified in OncoKB as likely neutral, arguing against therapeutic relevance in this context. *ATM* is involved in DNA damage response pathways, while the clinical and therapeutic relevance for the specific alteration *ATM R720H* in this case remains unclear. *RB1 F293L* was likely germline and clinically non-relevant. Despite the absence of clearly actionable mutations, the LION panel analysis provided clinically relevant insights. In particular, the increase in VAFs across multiple variants supports the presence of disease progression in a setting where imaging findings were inconclusive. Furthermore, liquid biopsy-based NGS enables minimally invasive longitudinal sampling, facilitating close monitoring of both disease dynamics and the evolving mutational landscape of this patient.

Taken together, these case studies demonstrate that targeted liquid biopsy-based LION panel analysis provides clinically relevant and potentially actionable information for treatment stratification in a real-world MTB setting. In particular, it enables the early detection of disease progression, supports longitudinal monitoring of MRD, and comprehensive characterization of tumor evolution, including subclonal dynamics and emerging resistance-associated mutations. These findings suggest that ctDNA-based profiling may complement routine clinical and radiological assessment and support earlier identification of molecular changes relevant for therapeutic decision-making.

To facilitate broader accessibility and implementation, the LION panel and pipeline will be made available as open resource, packaged in a containerized docker to ensure reproducible and user-friendly distribution across clinical and research settings.

Overall, while these results highlight the potential clinical utility of the LION panel, they currently represent a technical validation and proof-of-concept in a limited cohort. Further studies in larger MTB patient populations are required to confirm its clinical impact and to establish its role in routine precision oncology workflows.

### 4.1. Limitations of the study

This study has several limitations. First, the 109-gene LION panel may miss clinically relevant alterations outside the targeted gene regions. Second, the diverse study cohort reflects a real-world MTB population which enhances clinical relevance but limits generalizability. The small sample size and single-center, retrospective design may limit generalizability to other institutions and patient populations. Third, clinically relevant biomarkers such as tumor mutational burden (TMB), mismatch repair deficiency (MMR), homologous recombination deficiency (HRD) and microsatellite instability (MSI) cannot currently be assessed. Fourth, while reference cfDNA strengthened analytical validation, incomplete variant documentation by the reference material vendor may lead to overestimation of false positives, as some undocumented variants may be genuine but unreported (57). NPV and PPV estimates are limited by missing prevalence data (58,59). Finally, although the presented case studies support potential clinical utility, broader validation in larger, prospective cohorts is required to confirm the impact of LION panel analysis on clinical decision-making in MTB populations.

## 5 Conclusion

This study demonstrates that the LION panel, combined with the LION bioinformatic pipeline, represents a feasible and clinically informative approach for ctDNA-based molecular profiling in MTB settings. It enables minimally invasive longitudinal monitoring of tumor evolution and minimal residual disease, facilitates early detection of disease progression and treatment resistance, and supports the dynamic identification of potentially targetable variants.

Importantly, LION panel analysis complements established approaches such as tissue-based whole exome sequencing by capturing dynamic changes in the mutational landscape that are not accessible through static biopsies. This is particularly relevant in patients with advanced cancer, where repeated tissue sampling is often not feasible and timely molecular reassessment is critical for treatment adaptation.

Beyond its clinical performance, the LION panel and pipeline follow an open-resource strategy, providing a fully transparent, manufacturer-independent panel and bioinformatic pipeline packaged for reproducible deployment. This addresses an unmet clinical need for accessible and adaptable ctDNA workflows across diverse MTB settings.

In conclusion, the LION panel and pipeline support clinical decision-making in precision oncology by enabling longitudinal monitoring of tumor evolution, earlier detection of disease progression, and dynamic reassessment of targetable variants that are not captured by static tissue-based approaches.

## Supporting information

Supplementary Methos

## Abbreviations

cfDNA: cell-free DNA
CHIP: clonal hematopoiesis of indeterminate potential
ctDNA: circulating tumor DNA
ddPCR: digital droplet polymerase chain reaction
FFPE: formalin-fixed paraffin-embedded
gDNA: genomic DNA
NGS: next generation sequencing
MRD: minimal residual disease
MTB: molecular tumor board
VAF: variant allele frequency
WES: whole exome sequencing.

## Data accessibility

Code availability: All processing steps are available via a bash script at https://github.com/kunstner/UKSH_liquidbiopsy.

Raw data for reference line samples have been deposited to ENA under the accession number PRJEB113859.

All other data supporting the findings of this study are available within the article and its Supporting Information.

## Author contributions

S.F., A.K., M.F., L.R., I.H., F.S., S.D.-P., H.B., N.v.B. and E.D. contributed to the development of the study concept. M.F., N.v.B. and E.D. designed the NGS panel. N.G., T.G., F.A., J.S., M.H., S.M.J.F., M.O. provided samples. S.F., T.H., F.A., S.C.N. and E.D. performed the experiments. S.F., A.K., M.F., L.R., F.S. and E.D. developed the bioinformatic workflow. S.F., T.H., J.S., M.H. and S.M.J.F collected clinical data. S.F., A.K., M.F., S.S. and E.D. conducted the statistical analyses. S.F., A.K., M.F., T.H., N.G. and E.D. performed research. S.F., A.K. and E.D. drafted the manuscript. All authors contributed to data interpretation, revised the manuscript critically, and approved the final version.

## Acknowledgements

H.B. and A.K. acknowledge computational support from the OMICS compute cluster at the University of Lübeck.

E.D. and S.F. acknowledge Fikret Erdogan (Agilent Technologies) for valuable discussions and continuous support.

This work was supported by the DFG Research Infrastructure NGS_CC (project 407495230) as part of the Next Generation Sequencing Competence Network (project 423957469). NGS analyses were carried out at the Competence Centre for Genomic Analysis (Kiel).

The graphical abstract was created in https://BioRender.com.

## Supporting information

Supplementary Figure S1. Flowchart of the panel of normals filtering process

Supplementary Figure S2. Validation cohort characteristics and CONSORT flowchart of the study

Supplementary Figure S3. TapeStation quality control

Supplementary Figure S4. Reference cfDNA analysis comparing the sensitivity of Mutect2 and VarDict, the specificity and the per-sample counts of false positives and false negatives

Supplementary Figure S5. Mutational landscape of genes associated with clonal hematopoiesis of indeterminate potential (CHIP) in the validation cohort

Supplementary Figure S6. Longitudinal course of mutations detected by the LION panel in cfDNA samples of a breast cancer patient

Supplementary Figure S7. Longitudinal course of all mutations detected by the LION panel in cfDNA liquid biopsy samples of the melanoma patient

Supplementary Figure S8. Longitudinal course of mutations detected by the LION panel in cfDNA samples from the rectal cancer patient

### Supplementary Methods

Supplementary Table S1. Genes and associated hotspot mutations covered by the LION panel Supplementary Table S2. Individual sample-level sequencing depth metrics

Supplementary Table S3. Sequencing depth summary across sequencing runs

Supplementary Table S4. Analytical performance metrics for variant detection in the reference cfDNA samples, including sensitivity, specificity, accuracy, positive (PPV) and negative predictive value (NPV)

Supplementary Table S5. Comparison of analytical performance between Mutect2 and VarDict for SNP+InDel detection in reference cfDNA samples, including sensitivity, specificity, accuracy, positive (PPV) and negative predictive value (NPV)

Supplementary Table S6. Gene fusions identified in reference cfDNA samples by the LION panel using FACTERA

Supplementary Table S7. Sample collection, therapy response and treatment course of a breast cancer patient

Supplementary Table S8. Genomic DNA and gene fusion analysis from liquid biopsy samples of a breast cancer patient

Supplementary Table S9. Gene fusions identified in melanoma liquid biopsy cfDNA samples using the LION panel

Supplementary Table S10. Timeline of sample collection in relation to treatment course and therapy response in a rectal cancer patient

Supplementary Table S11. Genomic DNA and gene fusion analysis from a rectal cancer patient

## Declaration of generative AI and AI-assisted technologies in the writing process

The authors acknowledge the use of AI-assisted technologies (Deepl, ChatGPT) to improve the readability and language of this paper. The final manuscript was critically reviewed and edited by the authors, who take full responsibility for the content.

