## Supplementary Figure S1 for "A liquid biopsy-centered, pan-cancer, open next generation sequencing panel to support clinical decision-making (LION panel)"

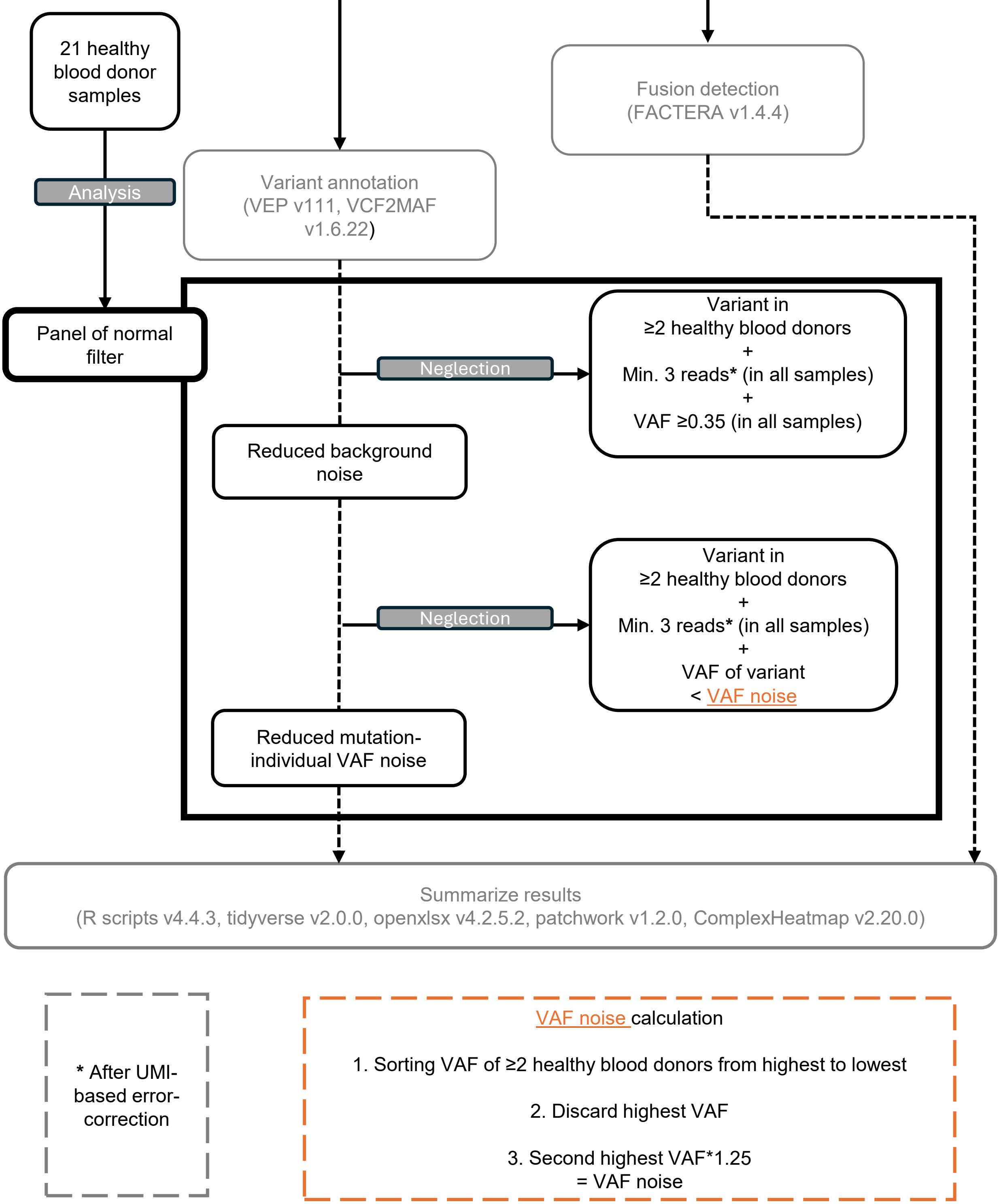


**Supplementary Figure S1. Flowchart of the panel of normals filtering process.**
The panel of normal includes two additional filter steps in the bioinformatic workflow (see Fig.1B) to remove annotated variants that occur in the general population and are not tumor-associated. The workflow was developed based on the analysis of 21 healthy blood donor samples. „in alle samples“= refers to all healthy blood donor samples in which the variant intended for filtering was detected.
Min., minimum; UMI, unique molecular identifier; VAF, variant allele frequency.
