## Supplementary Figure S2 for "A liquid biopsy-centered, pan-cancer, open next generation sequencing panel to support clinical decision-making (LION panel)"

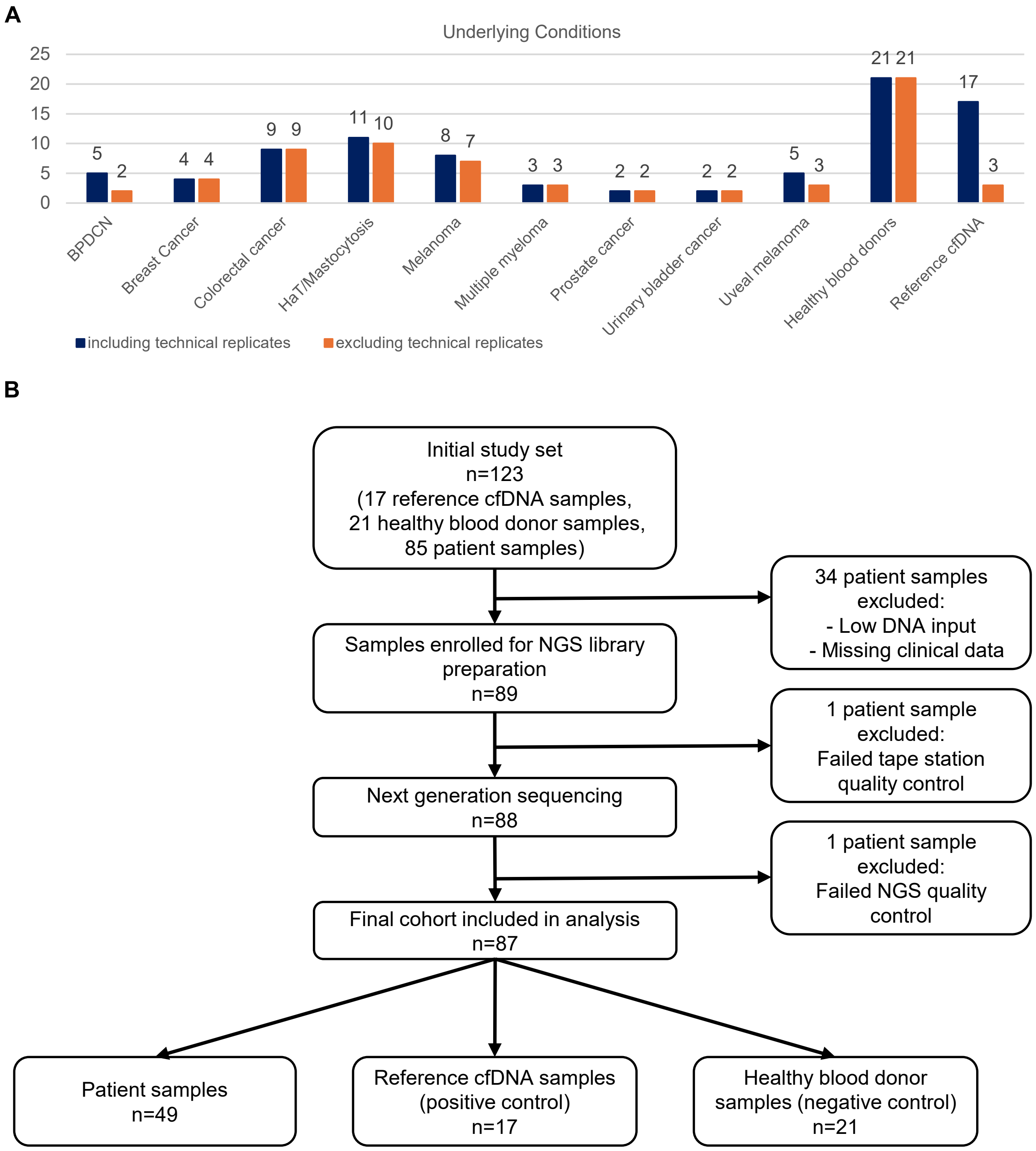


**Supplementary Figure S2. Validation cohort characteristics and CONSORT flowchart of the study.** (A) Validation cohort characteristics. Bars indicate numbers of samples including and excluding technical replicates. (B) CONSORT flowchart.

BPDCN, blastic plasmacytoid dendritic cell neoplasm; cfDNA, cell-free DNA; FFPE, formalin-fixed paraffin-embedded; HaT, hereditary alpha-tryptasemia; NGS, next generation sequencing.
