## Supplementary Figure S3 for "A liquid biopsy-centered, pan-cancer, open next generation sequencing panel to support clinical decision-making (LION panel)"

**Supplementary Figure S3. TapeStation quality control.** DNA quality control using TapeStation analysis following library preparation to assess DNA quantity and fragment size distribution prior sequencing. Each column represents one sample; A1 serves as the reference ladder. A total of 87 of 89 samples passed quality control, while two samples were excluded (*). The underlying condition is indicated for each sample; if two labels are shown, both diagnoses were present. BPDCN, blastic plasmacytoid dendritic cell neoplasm; cfDNA, cell-free DNA; FFPE, formalin-fixed paraffin-embedded; gDNA, genomic DNA; HaT, hereditary alpha-tryptasemia; NGS, next generation sequencing.

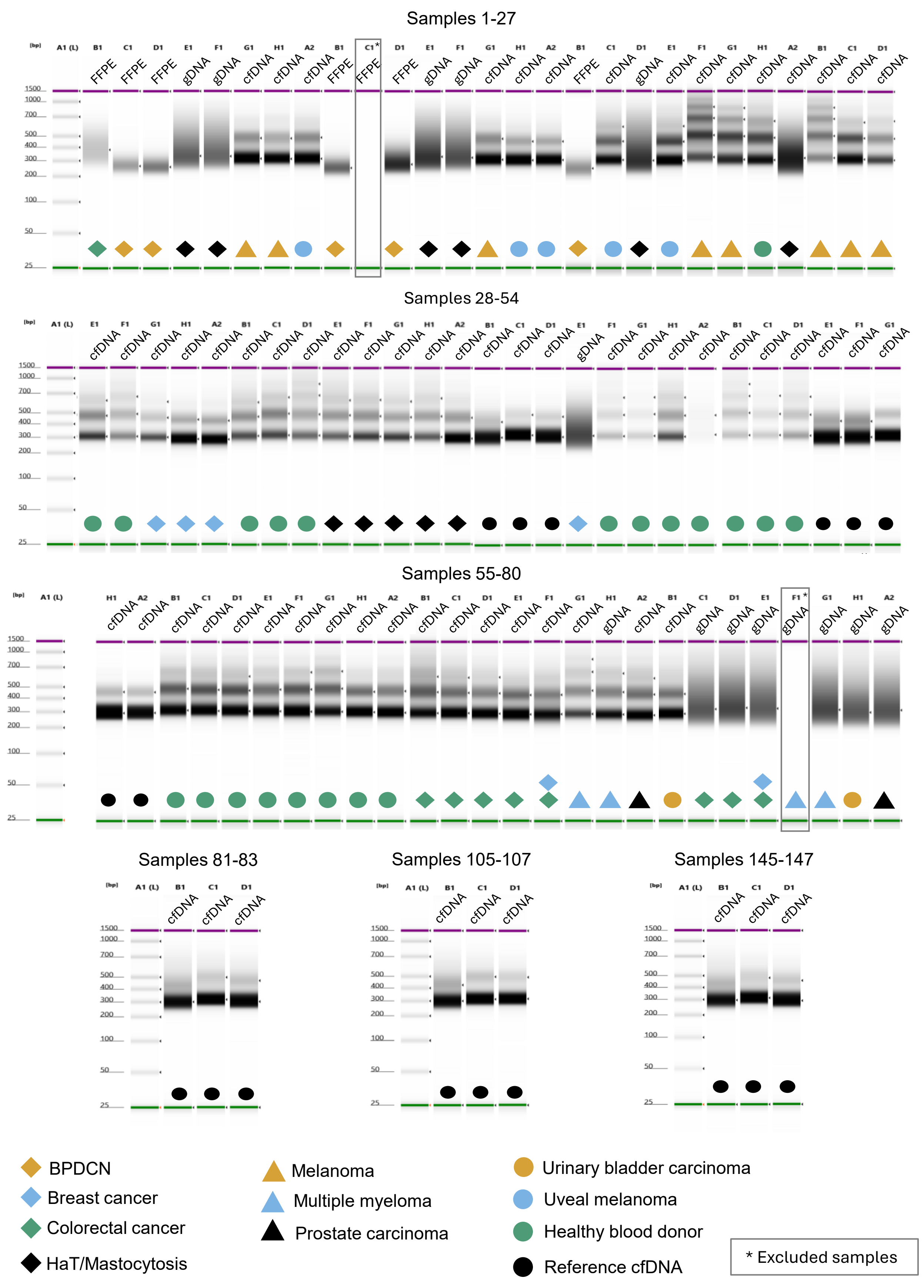

Samples 1-27
Samples 28-54
Samples 55-80
Samples 81-83
Samples 105-107
Samples 145-147
BPDCN
Breast cancer
Colorectal cancer
HaT/Mastocytosis
Melanoma
Multiple myeloma
Prostate carcinoma
Urinary bladder carcinoma
Uveal melanoma
Healthy blood donor
Reference cfDNA
* Excluded samples
