## Supplementary Figure S4 for "A liquid biopsy-centered, pan-cancer, open next generation sequencing panel to support clinical decision-making (LION panel)"

**Supplementary Figure S4. Reference cfDNA analysis comparing the sensitivity of Mutect2 and VarDict, the specificity and the per-sample counts of false positives and false negatives.**
(A) Comparison of pre-identified mutations detected tumor-uninformed in the reference cfDNA samples using the variant caller Mutect2 or Vardict at a 0.5% allele frequency threshold. (B) Summary of pre-identified mutations from 17 reference cfDNA samples that were not detected by the NGS panel. (C) Number of false positive mutations per reference cfDNA sample, categorized as SNPs, InDels or combined as SNPs+InDels. (D) Analytical specificity of variant detection across 17 reference cfDNA samples. Specificity values were consistently close to 1.0 for all variant types and calling tools. Minor differences are shown using a zoomed Y-axis (0.9998–1.000).
cfDNA, cell-free DNA; InDel, insertion/deletion; ng, nanogram; SNP, single nucleotide polymorphism; VAF, variant allele frequency.

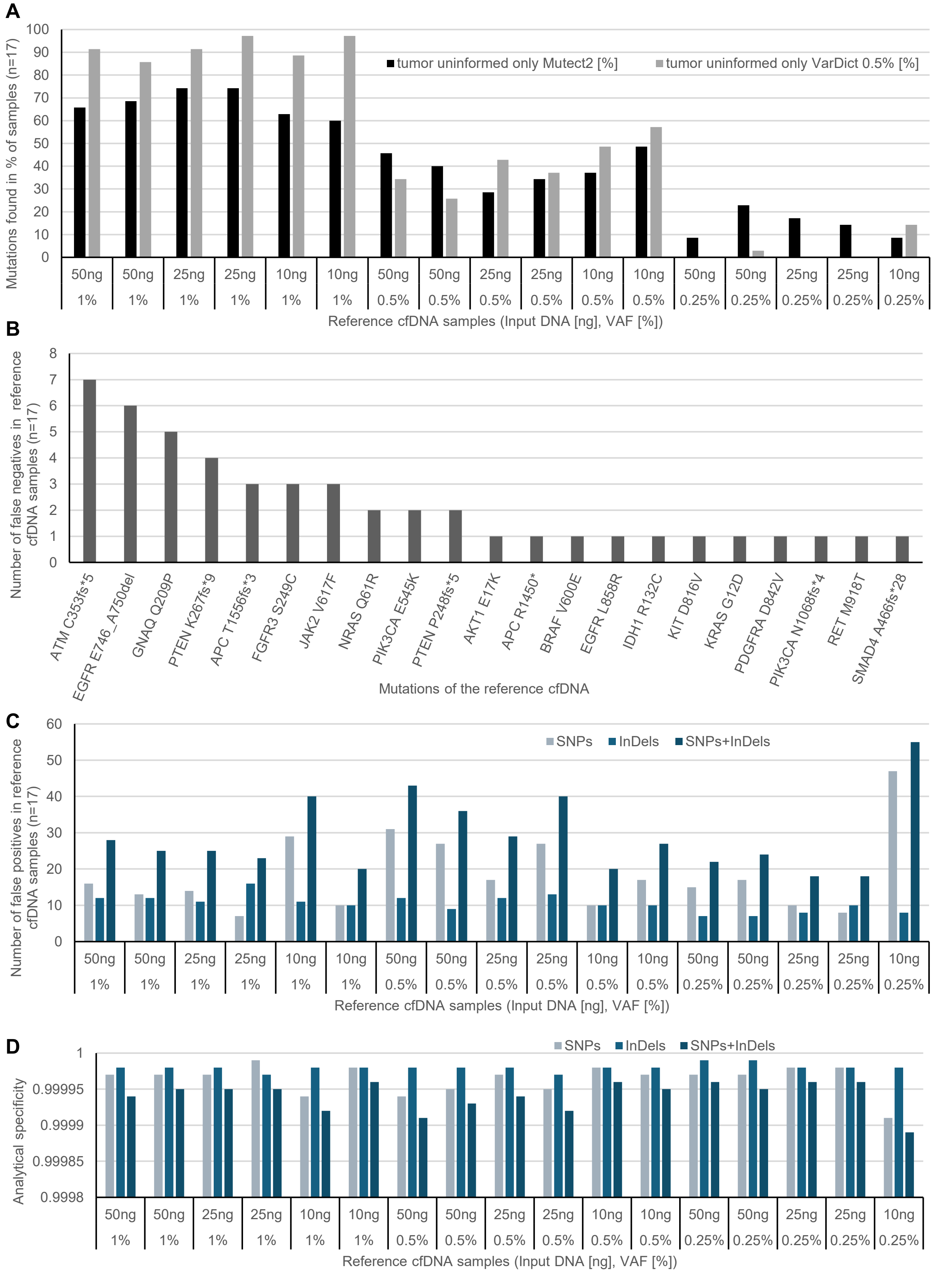

A
Mutations found in % of samples (n=17)
tumor uninformed only Mutect2 [%]
tumor uninformed only VarDict 0.5% [%]
Reference cfDNA samples (Input DNA [ng], VAF [%])
B
Number of false negatives in reference cfDNA samples (n=17)
ATM C353fs*5
EGFR E746_A750del
GNAQ Q209P
PTEN K267fs*9
APC T1556fs*3
FGFR3 S249C
JAK2 V617F
NRAS Q61R
PIK3CA E545K
PTEN P248fs*5
AKT1 E17K
APC R1450*
BRAF V600E
EGFR L858R
IDH1 R132C
KIT D816V
KRAS G12D
PDGFRA D842V
PIK3CA N1068fs*4
RET M918T
SMAD4 A466fs*28
Mutations of the reference cfDNA
C
Number of false positives in reference cfDNA samples (n=17)
SNPs
InDels
SNPs+InDels
Reference cfDNA samples (Input DNA [ng], VAF [%])
D
Analytical specificity
SNPs
InDels
SNPs+InDels
Reference cfDNA samples (Input DNA [ng], VAF [%])
