## Supplementary Figure S5 for "A liquid biopsy-centered, pan-cancer, open next generation sequencing panel to support clinical decision-making (LION panel)"

Supplementary Figure S5. Mutational landscape of genes associated with clonal hematopoiesis of indeterminate potential (CHIP) in the validation cohort.
Rows represent genes, columns represent individual samples. Colors indicate mutation and sample types.
ctDNA, circulating tumor DNA; FFPE, formalin-fixed paraffin-embedded; gDNA, genomic DNA.

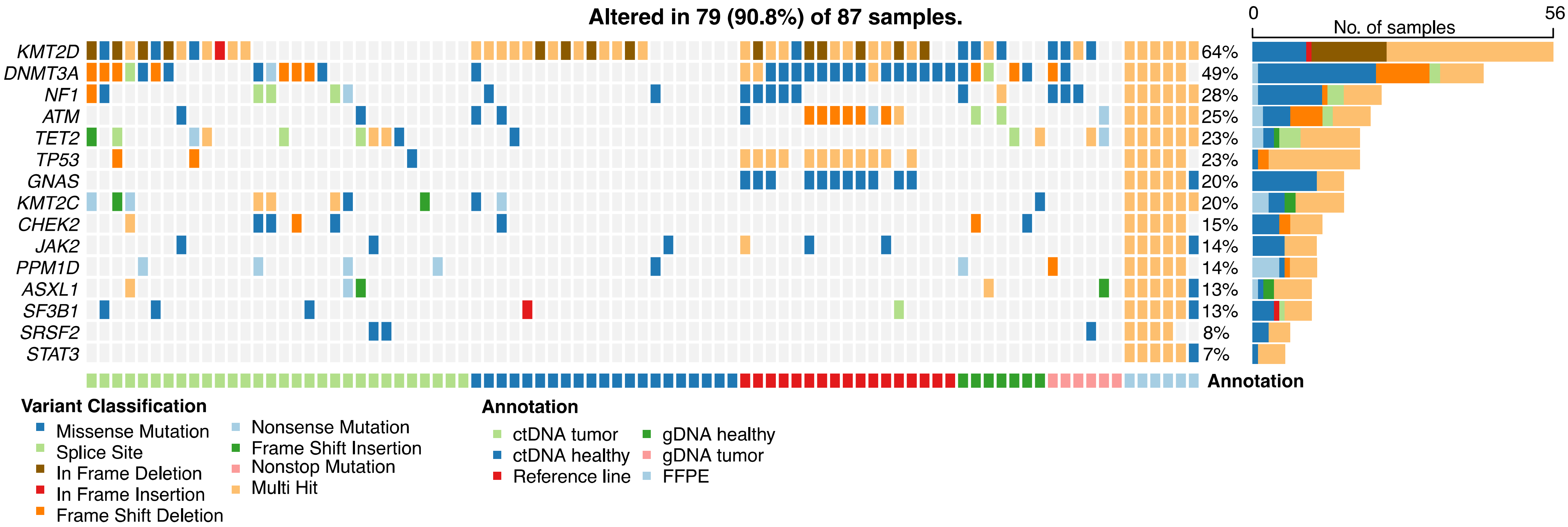
