## Supplementary Figure S6 for "A liquid biopsy-centered, pan-cancer, open next generation sequencing panel to support clinical decision-making (LION panel)"

**Supplementary Figure S6. Longitudinal course of mutations detected by the LION panel in cfDNA samples of a breast cancer patient.** Only mutations with a variant allele frequency (VAF) <45% across all samples are shown. The table lists each mutation and its corresponding VAF [%] at baseline, follow-up 1 and follow-up 2 sample.
cfDNA, cell-free DNA; VAF, variant allele frequency.

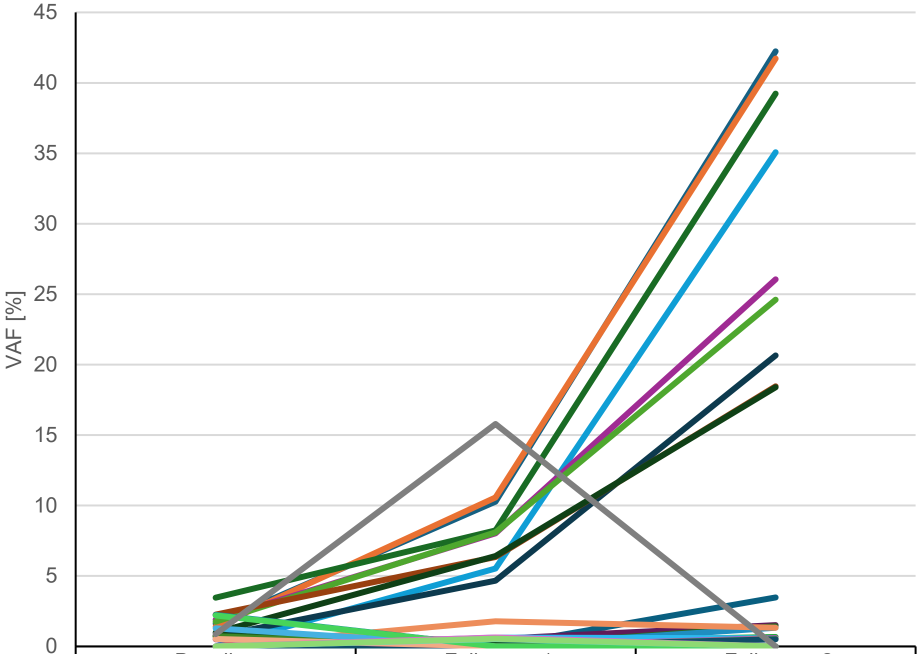


| | Baseline | Follow up 1 | Follow up 2 |
|---|---|---|---|
| NF1 X2657_splice | 1.6 | 10.3 | 42.2 |
| SMAD4 S178* | 1.4 | 10.6 | 41.7 |
| PIK3CA E545K | 3.5 | 8.2 | 39.2 |
| ABL1 Q40* | 0.0 | 5.5 | 35.1 |
| APC E1899K | 1.9 | 8.0 | 26.1 |
| APC E1989D | 1.8 | 8.1 | 24.6 |
| KMT2C S245F | 0.7 | 4.7 | 20.7 |
| NFE2L2 I295M | 2.3 | 6.3 | 18.5 |
| NFE2L2 Q494E | 1.0 | 6.4 | 18.4 |
| KEAP1 A184T | 0.0 | 0.0 | 3.5 |
| KMT2C I323M | 0.0 | 0.5 | 1.5 |
| DNMT3A E177* | 0.0 | 0.0 | 1.4 |
| LRP1B K3172T | 0.0 | 0.0 | 1.3 |
| TSC1 X876_splice | 0.0 | 1.8 | 1.3 |
| BRCA1 R1835Q | 0.0 | 0.3 | 0.7 |
| BRAF E26A | 0.0 | 0.6 | 0.6 |
| SOX9 E388K | 0.0 | 0.0 | 0.6 |
| KEAP1 E149* | 0.0 | 0.0 | 0.5 |
| EZH2 V637L | 0.0 | 0.0 | 0.5 |
| NOTCH2 E1223D | 0.7 | 0.0 | 0.0 |
| DNMT3A M761V | 0.8 | 0.4 | 0.0 |
| EZH2 V74G | 2.2 | 0.0 | 0.0 |
| KMT2C H3887Tfs*2 | 0.5 | 0.0 | 0.0 |
| DIS3 V417G | 0.7 | 0.0 | 0.0 |
| PPM1D Q26* | 1.3 | 0.0 | 0.0 |
| AXL M598R | 0.5 | 0.0 | 0.0 |
| ERBB2 c.2725+8_2725+9delinsTG | 2.2 | 0.0 | 0.0 |
| ARID1A Y154S | 0.0 | 0.5 | 0.0 |
| EGR1 S171L | 0.0 | 0.6 | 0.0 |
| ERBB2 c.2725+8A>T | 0.0 | 0.5 | 0.0 |
| UGT1A1 M137L | 0.9 | 15.8 | 0.0 |
