## Supplementary Figure S7 for "A liquid biopsy-centered, pan-cancer, open next generation sequencing panel to support clinical decision-making (LION panel)"

**Supplementary Figure S7. Longitudinal course of all mutations detected by the LION panel in cfDNA liquid biopsy samples of the melanoma patient.** A break in the Y-axis (between 12 and 42) is included for illustrative purposes only, as no data points fall within this range; allowing smaller values to be visualized clearly alongside a high-value outlier. The table lists each mutation and its corresponding variant allele frequency (VAF) in the baseline, follow-up 1, follow-up 2 and follow-up 3 samples. cfDNA, cell-free DNA; VAF, variant allele frequency.

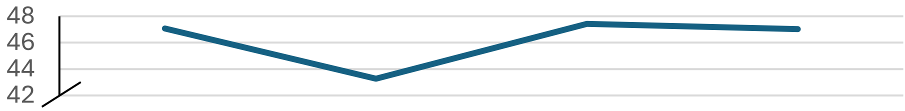


| | | | | |
|---|---|---|---|---|
| ALK p.V811M | 47.07 | 43.27 | 47.43 | 47.02 |

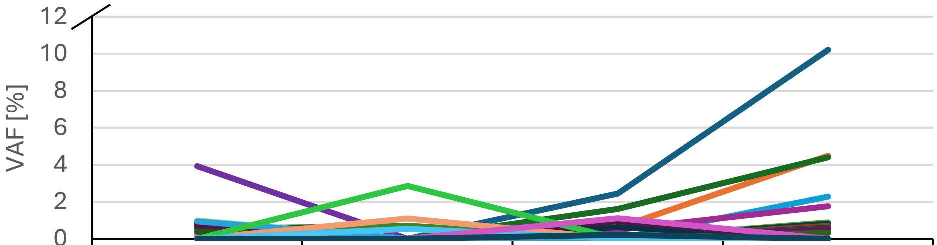


| | Baseline | Follow up 1 | Follow up 2 | Follow up 3 |
|---|---|---|---|---|
| BRAF p.V600E | 0.79 | 0.00 | 2.44 | 10.21 |
| MLH1 p.X702_splice | 0.00 | 0.00 | 0.60 | 4.49 |
| LRP1B p.G581V | 0.00 | 0.00 | 1.61 | 4.40 |
| PTEN p.S10_R11insSEIVS | 0.00 | 0.00 | 0.00 | 2.28 |
| PIK3R1 p.X582_splice | 0.00 | 0.00 | 0.49 | 1.76 |
| TCF7L2 p.S513A | 0.00 | 0.00 | 0.00 | 0.88 |
| KMT2D p.E473_E631del | 3.92 | 0.00 | 0.00 | 0.81 |
| RB1 p.T5P | 0.00 | 0.00 | 0.00 | 0.70 |
| TSC2 c.2220+7T>G | 0.00 | 0.00 | 0.00 | 0.60 |
| PIK3R1 p.K448Nfs*32 | 0.00 | 0.00 | 0.00 | 0.57 |
| ARID1A p.T118P | 0.00 | 0.00 | 0.00 | 0.53 |
| ARID1A p.A246G | 0.00 | 0.00 | 0.00 | 0.32 |
| SMO p.L23del | 0.59 | 0.00 | 0.00 | 0.00 |
| NOTCH1 p.C1345W | 0.62 | 0.00 | 0.00 | 0.00 |
| NOTCH1 p.X1158_splice | 0.65 | 0.00 | 0.00 | 0.00 |
| KMT2D p.P750S | 0.61 | 0.00 | 0.00 | 0.00 |
| KMT2D p.R487_S717del | 0.53 | 0.00 | 0.00 | 0.00 |
| KMT2D p.A596_E712del | 0.55 | 0.00 | 0.00 | 0.00 |
| KMT2D p.A464_E712del | 0.74 | 0.00 | 0.00 | 0.00 |
| KMT2D p.E455_E685del | 0.50 | 0.00 | 0.00 | 0.00 |
| NF1 c.61-3C>A | 0.55 | 0.67 | 0.00 | 0.00 |
| NOTCH2 p.P2300A | 0.12 | 0.00 | 0.00 | 0.00 |
| DNMT3A p.I780S | 0.34 | 0.00 | 0.00 | 0.00 |
| DNMT3A p.S638C | 0.28 | 0.71 | 0.00 | 0.00 |
| ABL1 p.S438A | 0.14 | 0.00 | 0.00 | 0.00 |
| NOTCH1 p.A1158G | 0.51 | 0.00 | 0.00 | 0.00 |
| CCND1 p.V52G | 0.27 | 0.00 | 0.00 | 0.00 |
| FLT3 p.S471P | 0.11 | 0.00 | 0.00 | 0.00 |
| ERBB2 p.G464R | 0.15 | 0.00 | 0.00 | 0.00 |
| ARID1A p.Y154S | 0.98 | 0.00 | 0.00 | 0.00 |
| LRP1B p.X1390_splice | 0.76 | 0.00 | 0.62 | 0.00 |
| NOTCH1 p.X619_splice | 0.66 | 0.00 | 0.00 | 0.00 |
| WT1 p.Q147P | 0.83 | 0.00 | 0.00 | 0.00 |
| KMT2D p.E587_E685del | 0.56 | 0.00 | 0.00 | 0.00 |
| ERBB2 c.2725+8A>T | 0.72 | 0.00 | 0.95 | 0.00 |
| KMT2D p.E455D | 0.39 | 0.00 | 0.00 | 0.00 |
| BRCA2 p.K3151N | 0.94 | 0.00 | 0.00 | 0.00 |
| BRAF p.S36A | 0.00 | 1.10 | 0.00 | 0.00 |
| TSC1 c.2626-4_2626-3delinsA | 0.00 | 2.86 | 0.00 | 0.00 |
| AXL p.X482_splice | 0.00 | 0.54 | 0.00 | 0.00 |
| TSC1 p.X876_splice | 0.00 | 0.00 | 1.11 | 0.00 |
| MTOR p.T2367S | 0.00 | 0.00 | 0.21 | 0.00 |
| MTOR p.M2366V | 0.00 | 0.00 | 0.22 | 0.00 |
