## Supplementary Figure S8 for "A liquid biopsy-centered, pan-cancer, open next generation sequencing panel to support clinical decision-making (LION panel)"

**Supplementary Figure S8. Longitudinal course of mutations detected by the LION panel in cfDNA samples from the rectal cancer patient.** Only mutations with a variant allele frequency (VAF) <40% across all samples are shown. The table lists each mutation and its corresponding VAF [%] in the baseline, follow-up 1 and follow-up 2 samples.
cfDNA, cell-free DNA; VAF, variant allele frequency.

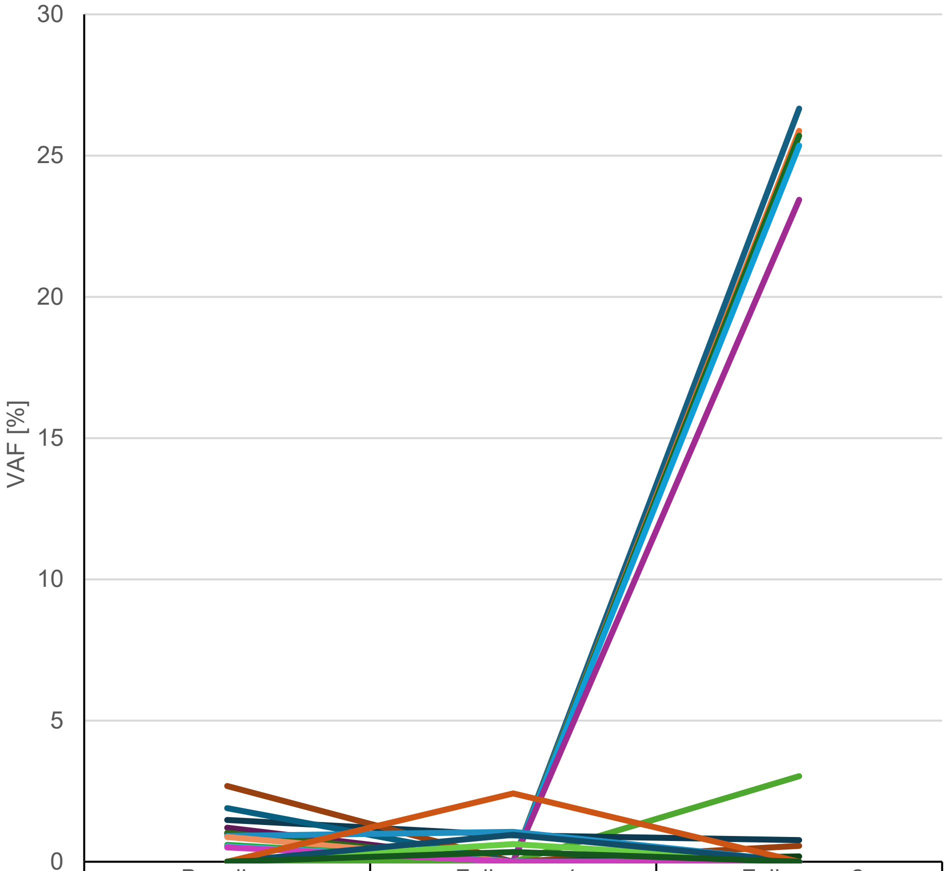


| | Baseline | Follow up 1 | Follow up 2 |
|---|---|---|---|
| PARP1 Q846R | 0.00 | 0.00 | 26.66 |
| PIK3R1 c.334+7A>G | 0.00 | 0.00 | 25.88 |
| MLH1 K618A | 0.00 | 0.00 | 25.70 |
| VHL G29R | 0.00 | 0.00 | 25.35 |
| ATM R720H | 0.00 | 0.00 | 23.43 |
| ASXL1 S664Rfs*6 | 0.00 | 0.00 | 3.03 |
| TET2 X1349_splice | 1.48 | 0.94 | 0.76 |
| TCF7L2 R488C | 2.68 | 0.00 | 0.57 |
| SMARCB1 K364del | 1.00 | 0.00 | 0.18 |
| ERBB3 T389K | 1.90 | 0.00 | 0.00 |
| APC Q999* | 1.20 | 0.00 | 0.00 |
| TP53 C135Afs*35 | 1.02 | 0.00 | 0.00 |
| DNMT3A W306Cfs*10 | 0.91 | 1.06 | 0.00 |
| APC Y1376* | 0.88 | 0.00 | 0.00 |
| KMT2C E2798Gfs*11 | 0.60 | 0.00 | 0.00 |
| KMT2C N2924K | 0.54 | 0.00 | 0.00 |
| KMT2D L617_P679del | 0.51 | 0.00 | 0.00 |
| KDR N143K | 0.00 | 0.62 | 0.00 |
| BRAF S36A | 0.00 | 0.95 | 0.00 |
| NF1 A188E | 0.00 | 2.42 | 0.00 |
| ASXL1 c.1085+7C>T | 0.00 | 0.34 | 0.00 |
