## Supplementary Methos for "A liquid biopsy-centered, pan-cancer, open next generation sequencing panel to support clinical decision-making (LION panel)"

### Supplementary Methods

#### Sample processing and DNA isolation

Peripheral blood samples from patients and healthy blood donors were collected in EDTA (Sarstedt, Nümbrecht, Germany) or cell free DNA-BCT® CE tubes (Streck, La Vista, USA). EDTA samples were processed within 4 hours and Streck samples within 7 days after collection. Plasma from EDTA tubes was separated by centrifugation with centrifuge 5810 R (Eppendorf SE, Hamburg, Germany) at 800 g for 10 min at room temperature (RT), followed by a second centrifugation in a falcon tube (Sarstedt, Nümbrecht, Germany) at 1000 g for 10 min. Plasma was aliquoted (2 ml) into cryotubes (Thermo Fisher Scientific, Waltham, USA) and stored at -80 °C. Plasma from Streck tubes was separated by centrifugation with centrifuge 5702 R (Eppendorf SE, Hamburg, Germany) at 2000 g for 10 min at RT, transferred to falcon tubes, centrifuged again under same conditions, aliquoted to FluidX 2D-coded cryogenic tubes (Azanta Life Sciences, Chelmsford, USA) and stored at -80 °C.

Buffy coat was collected from the same blood sample. For Streck tubes, after the first centrifugation step 1 to 1.5 ml of the buffy coat layer was transferred to a FluidX 2D-coded cryotube (Azanta Life Sciences, Chelmsford, USA), homogenized and stored at -80°C. For EDTA samples, an additional erythrocyte lysis step was performed using 3ml of G-DEX™ IIb RBC Lysis Buffer (iNtRON Biotechnology, Seongnam, South Korea) added to the transferred buffy coat layer. After vortexing with a mini vortexer (Thermo Fisher Scientific, Waltham, USA) and shaking for 10 min at RT, lysis was stopped with 11 ml of DPBS (Thermo Fisher Scientific, Waltham, USA). Samples were centrifuged with centrifuge 5810 R (Eppendorf SE, Hamburg, Germany) at 300 g for 10 min at 4 °C, the supernatant was discarded and the pellet resuspended in 1000 µl of PBS. After aliquoting in two 500 µl portions in Micro tube 1.5 ml Safe Seal (Sarstedt, Nümbrecht, Germany), the last centrifugation step was repeated with the mini centrifuge (SunLab®, Hamburg, Germany) and the supernatant was discarded, while the remaining pellet was stored at -80 °C.

Cell-free DNA (cfDNA) was isolated from 2 ml plasma from cancer patients or from 4 ml plasma from the healthy blood donors, either manually using QIAmp® Circulating Nucleic Acid Kit (Qiagen, Hilden, Germany) or automatically using the EZ2 Connect with the EZ 1 & 2 ccfDNA Kit (Qiagen, Hilden, Germany), according to the manufacturer’s instructions.

The genomic DNA (gDNA) from buffy coat was extracted using the QIAmp® DNA Mini Kit (Qiagen, Hilden, Germany) according to the manufacturer’s instructions. DNA from FFPE tissue of blastic plasmacytoid dendritic cell neoplasm (BPDCN) was extracted at an external pathology laboratory using the Maxwell® RSC DNA FFPE Kit (Promega, Madison, USA) and DNA from tumor resection of colorectal carcinoma was isolated from FFPE samples using AllPrep DNA/RNA FFPE Kit (Qiagen, Hilden, Germany), according to the manufacturer's instructions.

DNA concentration was measured with a Qubit 4 Fluorometer (Thermo Fisher Scientific, Waltham, USA) using either the Qubit™ ds DNA BR Assay Kit or Qubit™ 1X ds DNA HS Assay Kit (Thermo Fisher Scientific, Waltham, USA), according to the manufacturer’s instructions. Isolated DNA was stored at -20 °C until further use.

#### Library preparation and sequencing

Library preparation was performed using the SureSelect XT HS2 DNA Target Enrichment Kit (Agilent Technologies, Santa Clara, USA) according to the manufacturer’s instructions. The first 16 samples (Batch1; Table S2, Fig. S3) were processed manually before transitioning to automated library preparation using the Magnis Dx NGS Prep System (Agilent Technologies,

Santa Clara, USA). Whenever possible, approximately 50 ng of DNA input was used. For cfDNA, the maximum available DNA input was used, which could be 10 ng or less due to limited sample availability or low cfDNA concentration in plasma and a maximum of 50 ng for three samples. Genomic DNA (e.g. from buffy coat samples) underwent enzymatic fragmentation prior to end repair, while this step was skipped for cfDNA as it is already fragmented. DNA fragments were end-repaired, A-tailed and ligated to molecular-barcoded adapters (MBCs) containing unique molecular identifiers (UMIs). After bead-based purification using AMPure XP beads, libraries were amplified and indexed using eleven pre-capture PCR cycles, followed by purification. Pre-capture library quality and fragment size distribution were assessed using the D1000 ScreenTape kit on the Agilent 4150 TapeStation Instrument (Agilent Technologies, Santa Clara, USA). Following hybridization, captured DNA fragments were isolated using streptavidin-coated magnetic beads and amplified in 13 post-capture PCR cycles, followed by purification. The number of PCR cycles were adjusted according to the DNA input to ensure sufficient library material for sequencing while minimizing PCR-induced errors. Final library quality assessment included fragment size analysis and determination of the main peak (bp) using the High Sensitivity D1000 ScreenTape kit on the Agilent 4150 TapeStation Instrument (Agilent Technologies, Santa Clara, USA) as well as quantification of concentration using the Qubit™ ds DNA BR Assay Kiton the Qubit 4 Fluorometer (Thermo Fisher Scientific, Waltham, USA).

Libraries were multiplexed using 8 bp i5 and i7 dual indices incorporated during library preparation. For pooling, molar library concentrations were calculated using the formula c [nM] = c [ng/µl]/ (660 [g/mol] *main peak [bp]) *10^6. Libraries were diluted to 2 nM in a minimum volume of 10 µl and pooled at equimolar ratios. The first 16 samples were sequenced on a single lane of a NovaSeq 6000 SP flow cell (Illumina, San Diego, USA) using a pool volume of at least 28µl (2 µl of each 2nM library). The remaining samples were multiplexed with an average of 33 libraries per lane on a NovaSeq 6000 S4 flow cell (Illumina, San Diego, USA) with a pool volume of at least 40 µl. Paired-end sequencing (2x151bp) was performed in both cases. Sequencing targeted a mean on-target coverage after bioinformatic error-correction of approximately 500x for FFPE and 3000x for liquid biopsy samples, as recommended by the library kit manufacturer.
